# Long-term mental health outcomes after SARS-CoV-2 infection: prospective cohort study

**DOI:** 10.1101/2022.09.26.22280293

**Authors:** Yunhe Wang, Junqing Xie, Clemente Garcia, Daniel Prieto-Alhambra

## Abstract

Despite previous evidence from retrospective cohorts suggest that survivors of COVID-19 may be at increased risk of psychiatric sequelae, questions remain on the incidence and absolute risk of psychiatric outcomes, and on the potential protective effect of vaccination. Addressing these knowledge gaps will help public health and clinical service planning during the ongoing pandemic. Based on UK Biobank prospective data, we constructed a *SARS-CoV-2 infection cohort* including participants with a positive PCR test for SARS-CoV-2 between March 1, 2020 and September 30, 2021; a *contemporary control cohort* with no evidence of SARS-CoV-2, and a *historical control cohort* predating the COVID-19 pandemic. Additional control cohorts were constructed for benchmarking, including participants diagnosed with *other respiratory tract infection*, or with a *negative SARS-CoV-2 test*. We used propensity score weighting using predefined (clinically informed) and data-driven covariates to minimize confounding. We then estimated incidence rates and risk of first psychiatric disorders diagnosed by ICD-10 codes and psychotropic prescriptions after SARS-CoV-2 infection using cause-specific Cox models.

In this prospective cohort including 406,579 adults (224,681 women, 181,898 men; mean [SD] age 66.1 [8.4] years), 26,181 had a *SARS-CoV-2 infection*. Compared with *contemporary controls* (n=380,398), COVID-19 survivors had increased risks of subsequent psychiatric diagnoses (HR: 2.02, 95% CI 1.85-2.21; difference in incidence rate: 24.85, 95 CI 20.69-29.39 per 1000 person-years) and psychotropic prescriptions (HR: 1.61, 95% CI 1.48-1.75; difference in incidence rate: 21.77, 95% CI 16.59-27.54 per 1000 person-years). Regarding individual mental health related outcomes, the *SARS-CoV-2 infection* cohort showed an increased risk of psychotic disorders (2.26, 1.28-3.98), mood disorders (2.19, 1.92-2.50), anxiety disorders (2.08, 1.82-2.38), substance use disorders (1.59, 1.34-1.90), sleep disorders (1.95, 1.60-2.39); and prescriptions for antipsychotics (3.78, 2.74-5.21), antidepressants (1.55, 1.29-1.87), benzodiazepines (1.82, 1.58-2.11), and opioids (1.40, 1.26-1.55). Overall, the risk of any mental health outcome was increased with a HR of 1.58, 95% CI 1.47-1.70; and difference in incidence rate of 32.04, 25.76-38.81 per 1000 person-years. These results were consistent when comparing to a *historical control* cohort. Additionally, mental health risks were increased even further in participants who tested positive in hospital settings. Finally, participants who were fully vaccinated had a lower risk of mental health outcomes compared to those infected when unvaccinated or partially vaccinated. All observed risks of mental health outcomes were attenuated or even lower after *SARS-CoV-2 infection* compared with those with *other respiratory infections*, or with participants in the *test-negative control cohort*.

In this prospective cohort study, people who survived COVID-19 were at increased risk of psychiatric outcomes and related psychotropic medications. These risks were higher in those with more severe disease, treated in hospital settings, and were significantly reduced in fully vaccinated people. Of note, compared to participants with other respiratory infections or with only negative testing results, those infected with SARS-CoV-2 had an even lower risk of mental health outcomes, warranting further research into causation. The early identification and treatment of psychiatric disorders among survivors of COVID-19 should be a priority in the long-term management of COVID-19. Particular attention might be needed for those with severe (hospitalized) disease and those who were not fully vaccinated at the time of infection.

## Introduction

The continuing spread of coronavirus disease 2019 (COVID-19) caused by severe acute respiratory syndrome coronavirus 2 (SARS-CoV-2) remains a major public health concern and results in enormous disease burden, with more than 605.4 million cases and 6.5 million death registered worldwide as of September 9, 2022.^1^ Emerging evidence exists for the direct (through infection) and indirect (through change in environmental stressors and individual behaviors) effects of SARS-CoV-2 on the pulmonary and multiple extrapulmonary organs, including the metabolic, renal, and cardiovascular systems, during and beyond the acute phase of COVID-19 of any severity.^2-4^ Studies have also reported an increased risk of neurological and psychiatric disorders in individuals admitted to hospital for COVID-19 and those with mild or asymptomatic disease during 3 to 12 months after infection.^5-8^ However, these studies to date have been based on electronic health records (EHR) or registry data and only adjusted for several potential confounders. Notably, although EHR-based studies used different setting of negative control groups including individuals with influenza and other diseases, socioeconomic and lifestyle factors and public health interventions in the context of pandemic such as vaccination and quarantine measures that were associated with both SARS-CoV-2 infection and mental health conditions were only crudely measured or not available in these analyses,^5-8^ leading to substantive residual confounding and making causal interpretation of the findings challenging. Random well-controlled population-based cohort studies with detailed and robust recording of confounding factors and long-term follow up might be less subject to potential bias and confounding compared with registry data, and are in urgently need to improve the current understanding of the long-term psychiatric sequela of COVID-19.

In this study, we use data from UK Biobank to quantify the incidence and relative risk of psychiatric diagnoses and related psychotropic prescription in participants who had a positive test for SARS-CoV-2 during a maximal 12 months of follow-up after SARS-CoV-2 infection. We explored whether the association between COVID-19 and the subsequent psychiatric outcomes observed in previous EHR-based studies varied by test setting of SARS-CoV-2 infection and vaccination status.

## Methods

### Study design and participants

We used data from UK Biobank (https://www.ukbiobank.ac.uk/) to conduct this study. The UK Biobank is an ongoing community-based prospective cohort study, which recruited more than 500000 participants out of 9.2 million adults aged 40-70 years in the UK who were invited to participants (5.5% response rate), as detailed elsewhere.^9^ The baseline survey took place from 2006 to 2010 in 22 assessment centers. Overall, 503,317 participants provided informed written consent to take part in the study and be followed-up through linkage to health-related records. Polymerase-chain-reaction (PCR) based testing results for SARS-CoV-2 were obtained from Public Health England’s Second Generation Surveillance System (PHE-SGSS), a centralized microbiology database covering English clinical diagnostics laboratories that had been previously validated for COVID-19 research.^10^ Records of psychiatric diagnoses and relevant medications were obtained by linkage to Hospital Episode Statistics (HES) and general practitioners’ (GP) practices, and relevant confounding factors are available in UK Biobank.

### Cohort

We included UK Biobank participants from England who were still alive by March 1, 2020 (date of the first recorded COVID-19 case in the UK Biobank) to construct COVID-19 cohort. The COVID-19 cohort was defined as all individuals who had a positive result on a PCR test for SARS-CoV-2 confirmed between March 1, 2020 and September 30, 2021. The date when the first positive specimen sample was taken was set as the start of follow-up (*T*_*0*_) for infected cohort. Non-infected contemporary control group included individuals with no evidence of SARS-CoV-2 infection (those not in the infected cohort who had negative testing results or never tested). To ensure that contemporary control cohort had a similar the follow-up period as the infected cohort, a random index date during the same observational period (between March 1, 2020 and the end of follow-up) was assigned for the contemporary control cohort based on the distribution of *T*_*0*_ in the infected cohort, so that the proportion of participants followed up from a certain date was the same in both comparison cohorts. The end of follow-up period for both infected and contemporary control cohorts was September 30, 2021.

To further examine the associations between SARS-CoV-2 infection and outcomes related to mental health conditions compared to those unaffected by the COVID-19 pandemic, a historical control cohort was constructed by including participants from UK Biobank who were alive by March 1, 2018 and were not in the infected cohort. Similarly, the start of follow-up for participants in the historical control cohort was randomly assigned according to the distribution of *T*_*0*_ in infected cohort as *T*_*0*_ minus two years (730 days). The end of follow-up period for the historical control cohort was September 30, 2019.

To provide additional benchmarking for the incidence and risk of mental health outcomes, we constructed additional control cohorts including participants diagnosed with any respiratory tract infection including influenza, or participants with negative SARS-CoV-2 test results. Because of the atypically low incidence of influenza worldwide during the COVID-19 pandemic and its markedly different risk profile and disease severity compared with COVID-19,^11,12^ we did not specifically compare the risk of psychiatric sequelae of COVID-19 with these risk after influenza during the pandemic as previous studies did.^6,8^ Participants diagnosed with any respiratory tract infection between March 1, 2018 and March 1, 2020 were included. Any respiratory tract infection was defined as those with ICD-10 codes J00-06, J09-18, or J20-22, which were exactly consistent with the definition in previous EHR-based studies of psychiatric sequelae.^5,6^ Participants who tested negative for SARS-CoV-2 between March 1, 2020 and September 30, 2021, and were not in the infected cohort were included into test-negative cohort. Follow-up time of the two control cohorts was assigned to match the distribution of follow-up time in the infected cohort.

These cohorts were followed longitudinally to assess the incidence and risk of first or any (first or recurrent) psychiatric disorders and prescriptions for psychotropic medications during a maximal 12 months of follow-up after SARS-CoV-2 infection.

### Outcomes

The mental health related outcomes including psychiatric disorders and prescriptions for psychotropic medications were predefined based on prior knowledge and previous studies of COVID-19 psychiatric sequela.^5-8^ Psychiatric disorders were diagnosed based on ICD-10 codes (international classification of diseases, 10th revision), including psychotic disorders (F20-F29), mood disorders (F30-F39), anxiety disorders (F40-F48), substance use disorders (F12-F19), and sleep disorders (F51 and G47). We also investigated the major individual outcomes in each category separately. For example, the major components of mood disorders considered in this study included depressive episode (F32) and mania/bipolar affective disorder (F30-F31). Prescriptions for psychotropic medications were recorded in the UK Biobank database, including antidepressants, antipsychotics, benzodiazepines, and opioids. Detailed definitions of outcomes are provided in **Supplementary table 1**. We specified three composite outcomes of any psychiatric disorders (F20-F48), any prescription for psychotropic medications, and any psychiatric diagnosis or psychotropic prescription. Because psychiatric disorders tend to recur or relapse, we separately estimated the risk and incidence of the first incident mental health outcomes (eg, excluding participants with a history of the corresponding psychiatric disorders or psychotropic prescriptions in one year before the start of follow-up) and the risk and incidence of first or recurrent (prevalent) mental health outcomes (eg, including participants who had a diagnosis or record of related outcomes before the start of follow-up). The first incident mental health outcomes after the infection of SARS-CoV-2 were reported as the primary outcomes. Analyses of diagnostic subcategories and first or recurrent mental health outcomes are provided in the supplementary materials.

### Covariates

We used both predefined and data-driven covariates to adjust for the difference in baseline characteristics between comparison groups. Predefined covariates were selected based on prior knowledge, including a comprehensive set of established and suspected risk factors for COVID-19 and mental health conditions: age, sex, ethnicity, index of multiple deprivation^13^ (a summary contextual measure including seven aspects in crime, education, employment, health, housing, income and living environment used to represent socio-economic status), smoking status, physical activity, body mass index, systolic and diastolic blood pressure, estimated glomerular filtration rate, and hospital admissions. The battery of predefined covariates also included comorbidities identified using all clinical components of Charlson comorbidity index^14^: cancer, cerebrovascular disease, chronic kidney disease, chronic obstructive pulmonary disease, congestive heart failure, dementia, diabetes, HIV/AIDS, hemiplegia, myocardial infarction, liver disease, renal disease, peripheral vascular disease, and peptic ulcer disease.

To further reduce the risk of residual confounding and optimize adjustment of potential confounders, we included a list of data-driven clinical episodes diagnosed during patient hospitalization within one year before *T*_*0*_. We first classified 8,651 source ICD-10 diagnosis codes into 453 disease phenotype groups (DPGs) using a validated mapping algorithm (Phecode v1.2 ICD-10 map).^15^ We further selected DPG that occurred in more than 0.1% of participants into the adjustment after excluding rare DPGs that can hardly characterize a cohort and may lead to inconsistency in model estimation.^16,17^

### Statistical analyses

In the main analyses, we used propensity score (PS) weighting to control for difference in baseline characteristics between comparison groups (infected, contemporary control, and historical control). For each comparison pair, we built a multivariable logistic regression with Lasso L1 penalty to estimate the PS as the probability of belonging to exposure (infection) group and the probability of belonging in control group, using both predefined and data-driven DPGs. Inverse probability weights were calculated as one divided by the PS in the infected group and divided by one minus PS in control group. We also used PS matching as an alternative analytic approach in sensitivity analysis to verify the robustness of the results from PS weighting. Infected participants were matched 1:10 to the uninfected participants, with a caliper distance of 0.2 standard error of the logit of the PS and exact matching for *T*_*0*_. Any baseline characteristic with a absolute standardized mean difference (ASMD) between comparison groups lower than 0.1 was considered well balanced.^18^ We used cause-specific Cox proportional hazards regression models where death was considered as a competing risk to estimate hazard ratios (HRs) of mental health conditions between the infected and contemporary cohorts, and between the infected and historical cohorts, with the inverse probability weights applied when PS weighting was used. We also estimated the adjusted incidence rate per 1000 person-years in the infection cohort and the control cohort and the difference in incidence rate between comparison groups.

Regarding the risk of first composite mental health outcomes compared with contemporary control, we conducted subgroup analyses based on age, sex, BMI, IMD, and ethnicity. To assess whether contextual factors of COVID-19 such as quarantine measures may increase the risk of psychiatric sequelae, we undertook subgroup analyses by calendar period according to the timeline of UK government coronavirus lockdowns and measures (first stage: between January 2020 and January 2021; second stage: between January 2021 and September 2021). The first two national lockdowns came into force in England in the first stage and England entered third national lockdown on January 6, 2021.

To investigate whether the incidence and risk of mental health outcomes after SARS-COV-2 infection were affected by vaccination status (independent of vaccine types) and the test setting of COVID-19, we further categorized the SARS-COV-2 infection group as infection tested in hospital setting and community settings, and categorized the infection group as non-breakthrough infection (unvaccinated or partially 1-dose vaccinated at *T*_*0*_) and breakthrough infection (fully 2-dose vaccinated at *T*_*0*_). We compared these infection subgroups with control groups. Among infection group, we further compared the incidence and risk in those who tested positive in hospital setting with those tested positive in community settings, and compared these estimates in unvaccinated or partially vaccinated participants with fully vaccinated participants. We estimated the PS score and inverse probability weights for each infection subgroup using the same covariates and then estimated the incidence rate and risk of composite mental health outcomes separately in each subgroup using cause-specific Cox proportional hazards regression models.

To provide additional benchmarking for the incidence and risk of composite mental health outcomes, we additionally compare the incidence and risk of SARS-COV-2 infection group with the control groups of participants with any respiratory tract infection diagnosed before the pandemic. Comparisons were conducted using cause-specific Cox proportional hazards regression models balancing by weighting using a same set of covariates.

To assess the robustness of our main results of first composite mental health outcomes, we conducted multiple sensitivity analyses. Firstly, we used the 1:10 PS matching approach to construct contemporary and historical control with comparable characteristics. Secondly, we extended the look-back window for the data-driven clinical variables to up three years. Finally, we excluded participants with a history of the mental health related outcomes in the two years before the start of follow-up and repeat the analyses of the first incident outcomes.

Statistical significance was determined by a 95% confidence interval (CI) that excluded 1 for ratios and 0 for rate differences. All analyses and data visualizations were conducted using R statistical software (version 4.1).

## Results

Study design and the process of cohort construction are shown in **Figure 1**. The primary cohorts comprised 26,181 participants in the SARS-COV-2 infection group, 380,398 in the contemporary control group and 384,030 in the historical control group. Median follow-up in the SARS-COV-2 infection, contemporary control and historical control groups was 239, 255, and 242 days, respectively. Person years of follow-up in the three groups were 11,150, 193,379, and 206,059, respectively, altogether corresponding to a 410,588 person years of follow-up. The demographic and medical characteristics of SARS-CoV-2 infection, contemporary control, and historical control groups after weighting are shown in **Table 1** and characteristics of three groups before weighting are shown in **Supplementary Table 2**.

**Figure 1.**
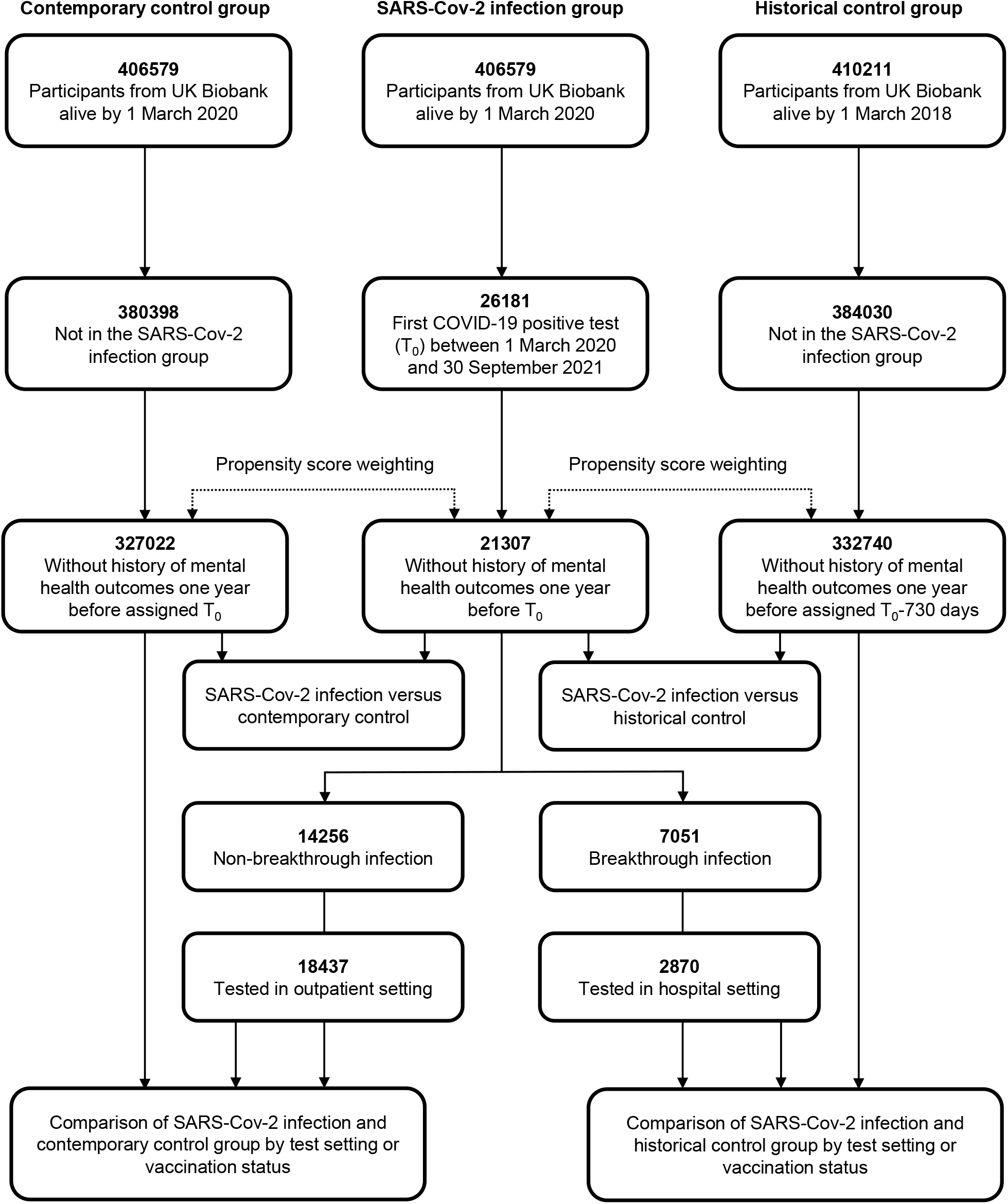
Flowchart of study design and cohort construction.

**Table 1.** Demographic and medical characteristics of SARS-CoV-2 infection, contemporary control, and historical control cohorts after weighting.

| Characteristics | SARS-CoV-2<br>infection<br>(n=26181) | Contemporary<br>control<br>(n=380398) | Historical<br>control<br>(n=384030) | ASMD between infection<br>and contemporary control* | ASMD between infection<br>and historical control* |
| --- | --- | --- | --- | --- | --- |
| Age, mean (sd) | 68.5 (8.4) | 68.6 (8.1) | 67.6 (8.2) | 0.01 | 0.03 |
| Sex, male (%) | 11860 (45.3) | 170038 (44.7) | 175559 (44.6) | 0.01 | 0.01 |
| Ethnicity, White (%) | 24191 (92.4) | 354151 (93.1) | 366778 (93.2) | 0.03 | 0.03 |
| Index of Multiple Deprivation, mean (sd) | 18.2 (13.7) | 17.5 (13.9) | 17.4 (13.8) | 0.05 | 0.07 |
| Body Mass Index, mean (sd) | 27.4 (4.6) | 27.4 (4.7) | 27.3 (4.7) | 0.02 | 0.03 |
| Current smoker (%) | 2723 (10.4) | 36899 (9.7) | 38465 (9.8) | 0.02 | 0.03 |
| Current drinker (%) | 23877 (91.2) | 349586 (91.9) | 361646 (91.9) | 0.03 | 0.03 |
| Physical activity, high level (%)<sup>#</sup> | 8509 (32.5) | 124770 (32.8) | 129879 (33.0) | 0.01 | 0.02 |
| Vaccination status, fully-vaccinated (%) | 10708 (40.9) | 148736 (39.1) | NA | 0.04 | NA |
| Medications (%)<sup>†</sup> |  |  |  |  |  |
| Lipid lowering drugs | 9477 (36.2) | 135041 (35.5) | 134186 (34.1) | 0.02 | 0.02 |
| RAS inhibitors | 6362 (24.3) | 90915 (23.9) | 91464 (23.3) | 0.01 | 0.02 |
| Other anti-hypertensives | 2906 (11.1) | 41083 (10.8) | 42070 (10.7) | 0.01 | 0.02 |
| Anticoagulants | 1152 (4.4) | 16357 (4.3) | 15285 (3.9) | 0.01 | 0.01 |
| Antiplatelet drugs | 3063 (11.7) | 42985 (11.3) | 42680 (10.8) | 0.01 | 0.03 |
| Proton pump inhibitors | 7880 (30.1) | 111837 (29.4) | 109661 (27.9) | 0.02 | 0.04 |
| Diabetes medicines | 1885 (7.2) | 25867 (6.8) | 26279 (6.7) | 0.02 | 0.03 |
| Systemic glucocorticoids | 1440 (5.5) | 20161 (5.3) | 24673 (6.3) | 0.01 | 0.02 |
| Immunosuppressants | 340 (1.3) | 4564 (1.2) | 4763 (1.2) | 0.01 | 0.01 |
| Antineoplastic agents | 26 (0.1) | 380 (0.1) | 406 (0.1) | 0.01 | 0.01 |
| Coexisting conditions (%)<sup>†</sup> |  |  |  |  |  |
| Acquired immunodeficiency syndrome | 27 (0.1) | 394 (0.1) | 385 (0.1) | 0.01 | 0.01 |
| Cancer | 2801 (10.7) | 40322 (10.6) | 41893 (10.7) | 0.01 | 0.01 |
| Cerebrovascular disease | 628 (2.4) | 8749 (2.3) | 8344 (2.1) | 0.01 | 0.01 |
| Chronic obstructive pulmonary disease | 4503 (17.2) | 63526 (16.7) | 63757 (16.2) | 0.01 | 0.02 |
| Chronic kidney disease | 628 (5.5) | 20161 (5.3) | 19503 (5.0) | 0.01 | 0.01 |
| Congestive heart failure | 419 (1.6) | 5706 (1.5) | 5178 (1.3) | 0.01 | 0.01 |
| Dementia | 236 (0.9) | 3423 (0.9) | 2791 (0.7) | 0.01 | 0.01 |
| Diabetes (uncomplicated) | 2670 (10.2) | 36518 (9.6) | 36047 (9.2) | 0.02 | 0.04 |
| Diabetes (end-organ damage) | 838 (3.2) | 11792 (3.1) | 11729 (3.0) | 0.01 | 0.01 |
| Hemiplegia | 28 (0.1) | 407 (0.1) | 424 (0.1) | 0.01 | 0.01 |
| Liver disease | 209 (0.8) | 2663 (0.7) | 2780 (0.7) | 0.01 | 0.01 |
| Peptic ulcer | 628 (2.4) | 9129 (2.4) | 9142 (2.3) | 0.01 | 0.01 |
| Rheumatoid arthritis | 785 (3.0) | 11411 (3.0) | 10757 (2.7) | 0.01 | 0.01 |
| Blood pressure, mean (sd), mm Hg |  |  |  |  |  |
| Systolic blood pressure | 139.3 (19.4) | 139.4 (19.2) | 139.2 (19.2) | 0.01 | 0.01 |
| Diastolic blood pressure | 82.2 (10.6) | 82.1 (10.5) | 82.0 (10.5) | 0.01 | 0.01 |
| Hospital admissions, mean (sd) <sup>†</sup> | 0.51 (1.69) | 0.42 (3.42) | 0.44 (2.40) | 0.03 | 0.06 |

Abbreviations: SD, standard deviation; MET, metabolic equivalent of task; ASMD, absolute standardized mean difference.
<sup>#</sup> Physical activity status was measured by the International Physical Activity Questionnaire (IPAQ).
<sup>†</sup>Data collected within past one year of T<sub>0</sub> from primary care records.
\*ASMD ≤0.10 is considered good balance between comparison groups.

### Risk of mental health outcomes after SARS-COV-2 infection

#### COVID-19 group versus contemporary control

Before weighting, participants in the infection group were younger (mean age: 66.0 years vs 68.8 years), less likely from the White ethnic group (84.6% vs 93.7%), more socioeconomically deprived (mean IMD: 20.5 vs 17.3), and more physically obese (mean BMI: 28.1 vs 27.3), compared with contemporary controls (**Supplementary Table 2**). After weighting, all characteristics were well balanced between two comparison groups (ASMD <0.1) and index dates were fully aligned (**Table 1**). The incidence and risk of the first incident psychiatric diagnoses and prescriptions for psychotropic medications in these groups are provided in **Figure 2**. The incidence and risk of first or recurrent mental health outcomes are shown in **Supplementary Figure 2**. The incidence and risk of composite mental health outcomes are provided in **Figure 3**.

**Figure 2.**
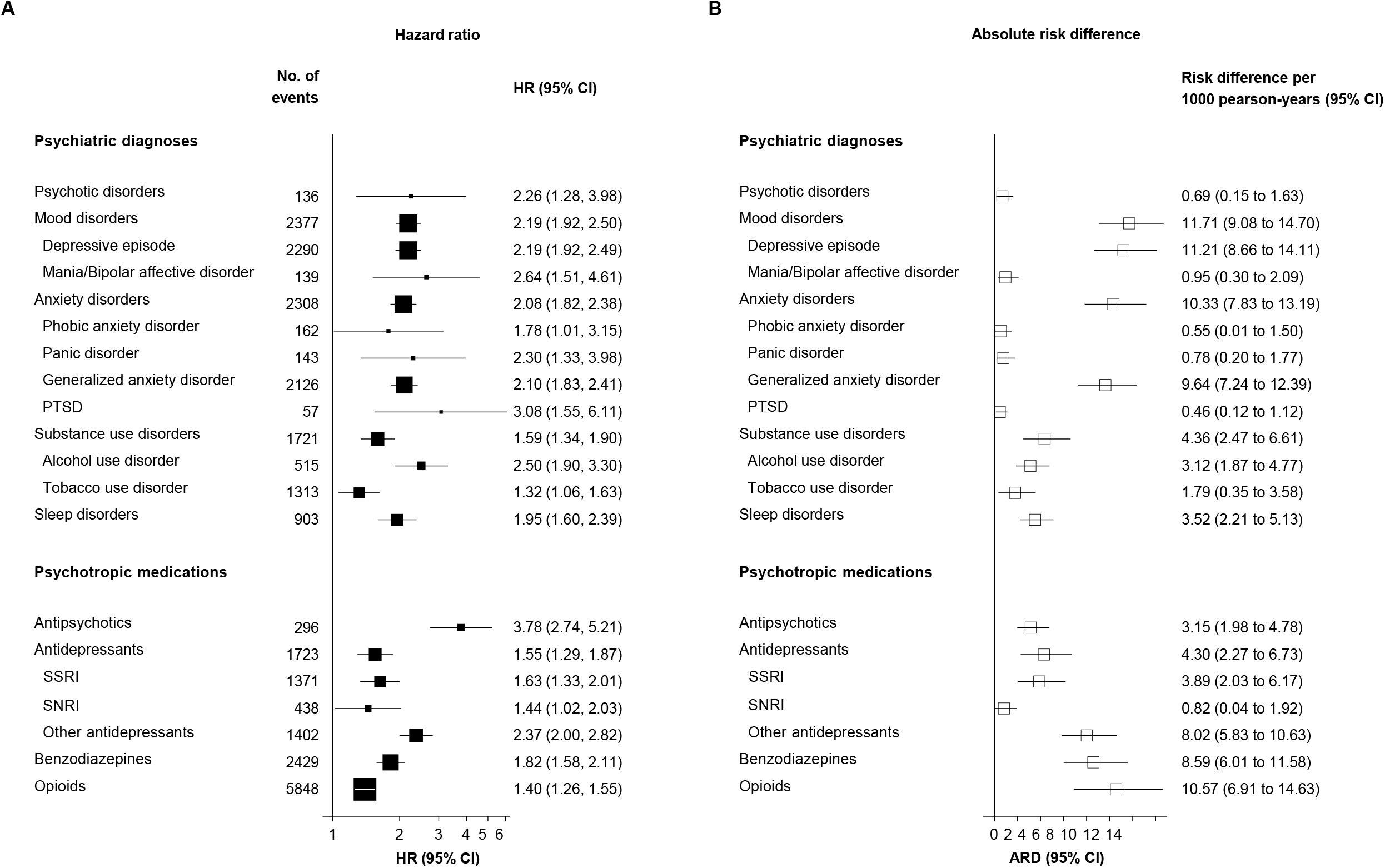
Risks of first psychiatric diagnoses and prescriptions for psychotropic medications after SARS-Cov-2 infection compared with the contemporary control group. Mental health outcomes were ascertained after the SARS-CoV-2 infection until the end of follow-up. Hazard ratios were adjusted for predefined and data-driven covariates. SSRI=selective serotonin reuptake inhibitor; SNRI=serotonin-noradrenaline reuptake inhibitor; PTSD=post-traumatic stress disorder

**Figure 3.**
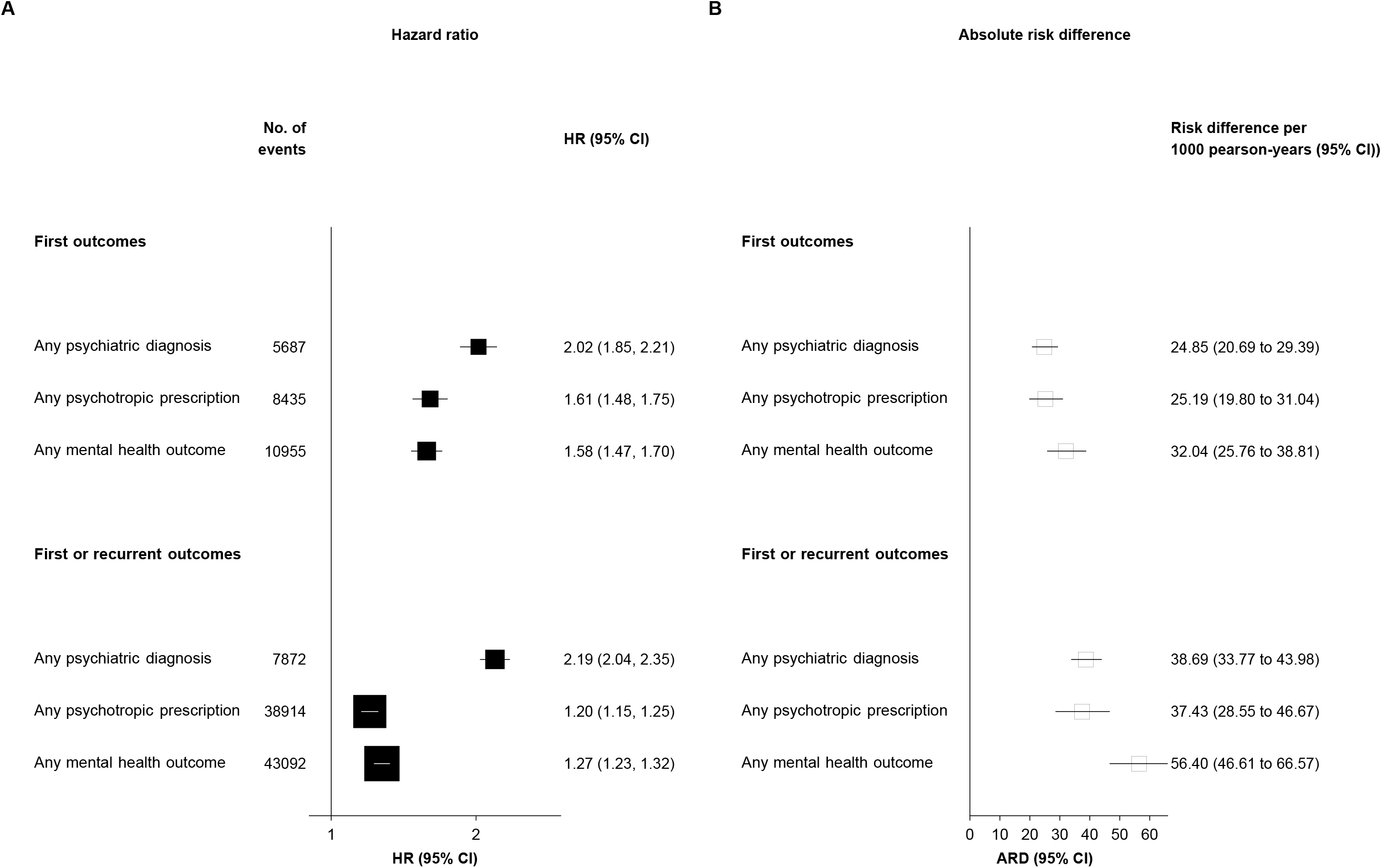
Risks of composite mental health outcomes after SARS-Cov-2 infection compared with the contemporary control group. Mental health outcomes were ascertained after the SARS-CoV-2 infection until the end of follow-up. Hazard ratios were adjusted for predefined and data-driven covariates.

##### Psychotic, mood, or anxiety disorders

Compared with contemporary control group, participants in SARS-CoV-2 infection group were at an increased risk of the first psychotic disorders (HR: 2.26, 95 CI 1.28-3.98; difference in incidence rate: 0.69, 95% CI 0.15-1.63 per 1000 person-years), mood disorders (2.19, 1.92-2.50; 11.71, 9.08-14.70 per 1000 person-years), and anxiety disorders (2.08, 1.82-2.38; 10.33, 7.83-13.19 per 1000 person-years) after COVID-19. There was an increased risk of individual diagnoses of mood disorders including depressive episode (2.19, 1.92-2.49; 11.21, 8.66-14.11 per 1000 person-years) and mania/bipolar affective disorder (2.64, 1.51-4.61; 0.95, 0.30-2.09 per 1000 person-years), and anxiety disorders including phobic anxiety disorder (1.78, 1.01-3.15; 0.55, 0.01-1.50 per 1000 person-years), panic disorder (2.30, 1.33-3.98; 0.78, 0.20-1.77 per 1000 person-years), generalized anxiety disorder (2.10, 1.83-2.41; 9.64, 7.24-12.39 per 1000 person-years), and post-traumatic stress disorder (3.08, 1.55-6.11; 0.46, 0.12-1.12 per 1000 person-years).

##### Antipsychotics, antidepressants, or benzodiazepines

Coupled with the increased of psychiatric disorders, there were increased risks of the first prescriptions for antipsychotics (3.78, 2.74-5.21; 3.15, 1.98-4.78 per 1000 person-years), antidepressants (1.55, 1.29-1.87; 4.30, 2.27-6.73 per 1000 person-years), and benzodiazepines (1.82, 1.58-2.11; 8.59, 6.01-11.58 per 1000 person-years). The risk of prescriptions for subtypes of antidepressants including SSRI, SNRI, and others were also increased.

##### Opioids

The risk of the first opioid prescriptions was increased (1.40, 1.26-1.55; 10.57, 6.91-14.63 per 1000 person-years).

##### Substance use disorders

The risk of the first substance use disorders was increased (1.59, 1.34-1.90; 4.36, 2.47-6.61 per 1000 person-years). For individual outcomes, there were increased risks of of alcohol use disorder (1.32, 1.06-1.63; 3.12, 1.87-4.77 per 1000 person-years) and tobacco use disorder (1.32, 1.06-1.63; 1.79, 0.35-3.58 per 1000 person-years).

##### Sleep disorders

The risk of the first sleep disorders was increased (1.95, 1.60-2.39; 3.52, 2.21-5.13 per 1000 person-years).

##### Composite outcomes

Compared with contemporary control group, participants in SARS-CoV-2 infection group were at an increased risk of any first psychiatric diagnoses (2.02, 1.85-2.21; 24.85, 20.69-29.39 per 1000 person-years), any first prescriptions for psychotropic medications (1.61, 1.48-1.75; 25.19, 19.80-31.04 per 1000 person-years), and any first mental health related outcomes (1.58, 1.47-1.70; 32.04, 25.76-38.81 per 1000 person-years) after COVID-19. **Figure 4** shows the Kaplan-Meier curves for the first composite mental health outcomes.

**Figure 4.**
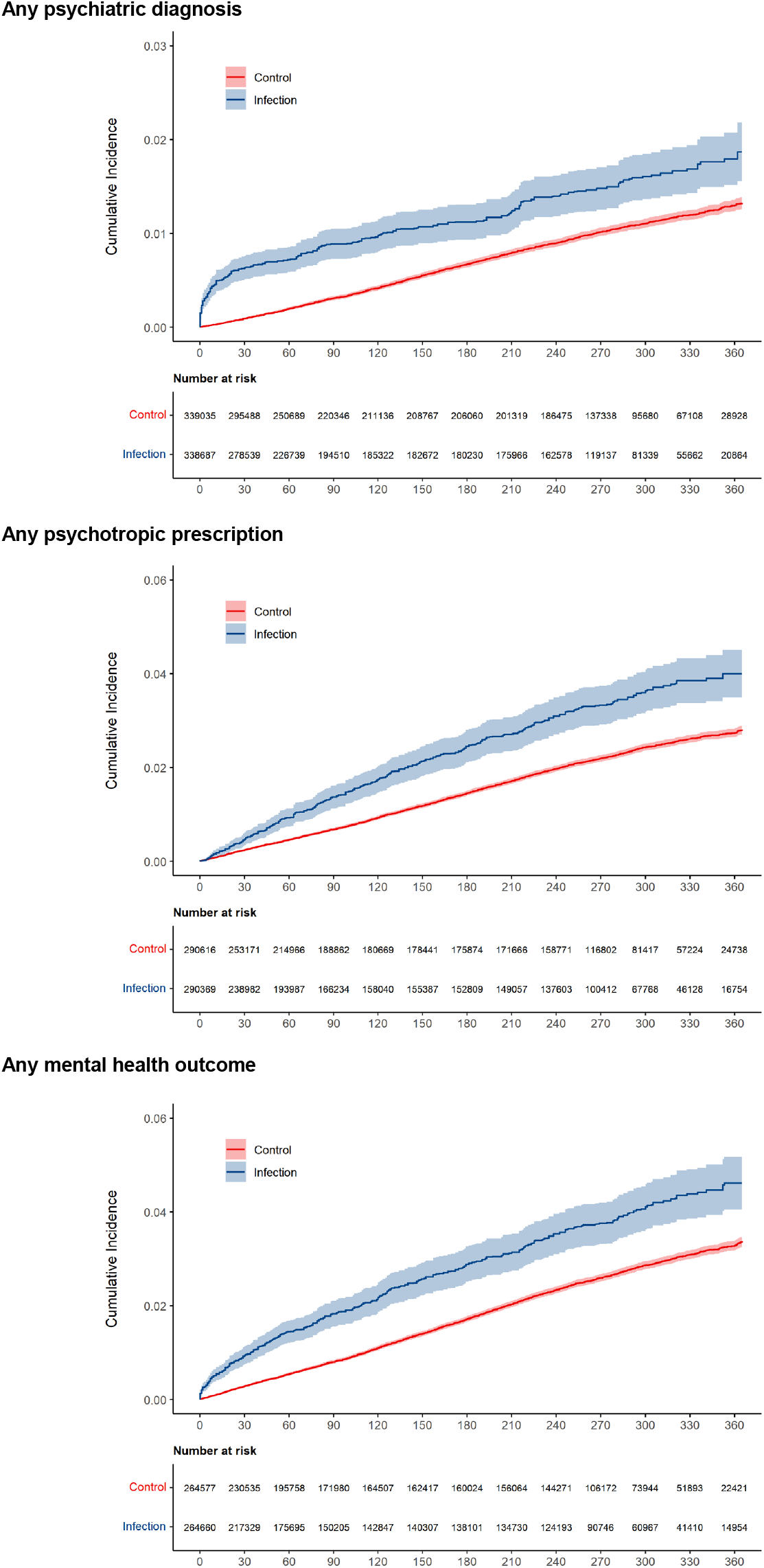
Cumulative incidence curves the first mental health outcomes after SARS-Cov-2 infection compared with those in the contemporary control group. Mental health outcomes were ascertained after the SARS-CoV-2 infection until the end of follow-up. Hazard ratios were adjusted for predefined and data-driven covariates

##### First or recurrent outcomes

Overall, the risks of first or recurrent psychiatric diagnoses (2.19, 2.04-2.35; 38.69, 33.77-43.98 per 1000 person-years) and prescriptions for psychotropic medications (1.20, 1.15-1.25; 37.43, 28.55-46.67 per 1000 person-years) were increased as was the risk of any first or recurrent mental health outcomes (1.27, 1.23-1.32; 56.40, 46.61-66.57 per 1000 person-years).

##### Subgroup analyses

The risks of incident composite mental health outcomes were consistently increased in all subgroups based on age, BMI, IMD, sex, ethnicity, and calendar period (**Figure 5**).

**Figure 5.**
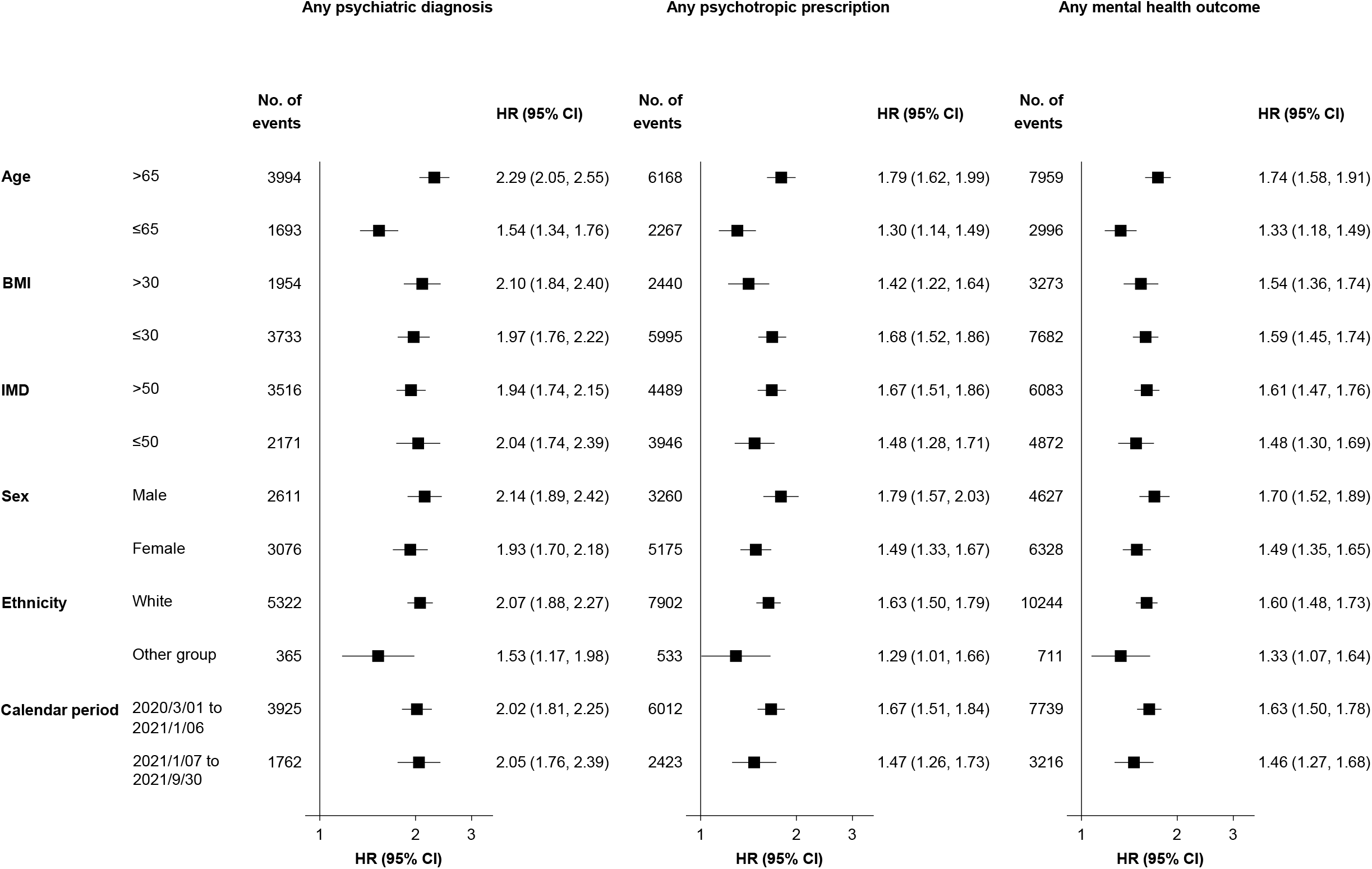
Subgroup analyses of the risks of the first mental health outcomes after SARS-Cov-2 infection compared with those in the contemporary control group. Mental health outcomes were ascertained after the SARS-CoV-2 infection until the end of follow-up. Hazard ratios were adjusted for predefined and data-driven covariates. BMI=body mass index; IMD=index of multiple deprivation.

#### COVID-19 group versus contemporary control by vaccination status or test setting

We further conducted analyses in mutually exclusive groups by test setting of infection or vaccination status of participants infected with SARS-CoV-2. Among SARS-CoV-2 infection group without history mental health outcomes one year before follow-up, 2870 participants were tested positive in hospital setting and 18437 were tested positive in community setting. 14256 participants were unvaccinated or partially vaccinated, and 7051 participants were fully vaccinated when tested positive. Assessment of covariate balance after PS weighting suggested that demographic and medical characteristics of these groups were well balanced. Compared with contemporary control group, the risks of both incident and prevalent mental health outcomes were increased in participants with non-breakthrough infection, and not significant or negative association were observed between breakthrough infection and mental health outcomes (**Figure 6**). Compared with non-breakthrough infection, breakthrough infection was associated with lower risks of first mental health outcomes (any mental health outcome: HR 0.41, 0.30-0.56) (**Supplementary Table 3**). Compared with contemporary control group, the risks of first or prevalent mental health outcomes in the infection group were increased in those tested positive in community setting and were highest in those tested positive in hospital setting (**Figure 7**). Compared with those tested positive in community setting, those tested in hospital setting were associated with increased risks of first mental health outcomes (any mental health outcome, HR: 1.96, 1.64-2.34) (**Supplementary Table 4**).

**Figure 6.**
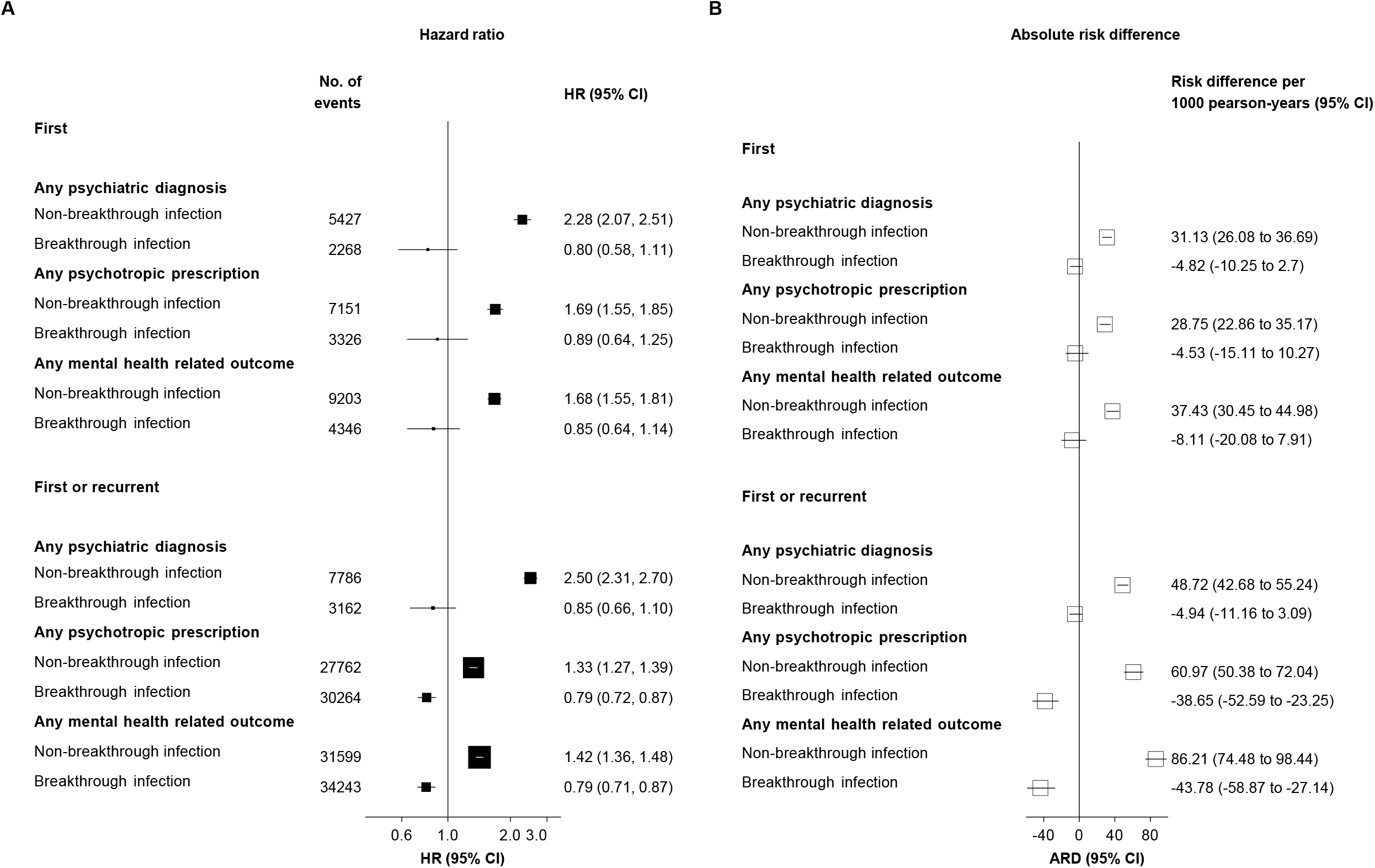
Risks of psychiatric diagnoses and prescriptions for psychotropic medications by vaccination status of participants in SARS-Cov-2 infection group compared with the contemporary control group. Mental health outcomes were ascertained after the SARS-CoV-2 infection until the end of follow-up. Hazard ratios were adjusted for predefined and data-driven covariates.

**Figure 7.**
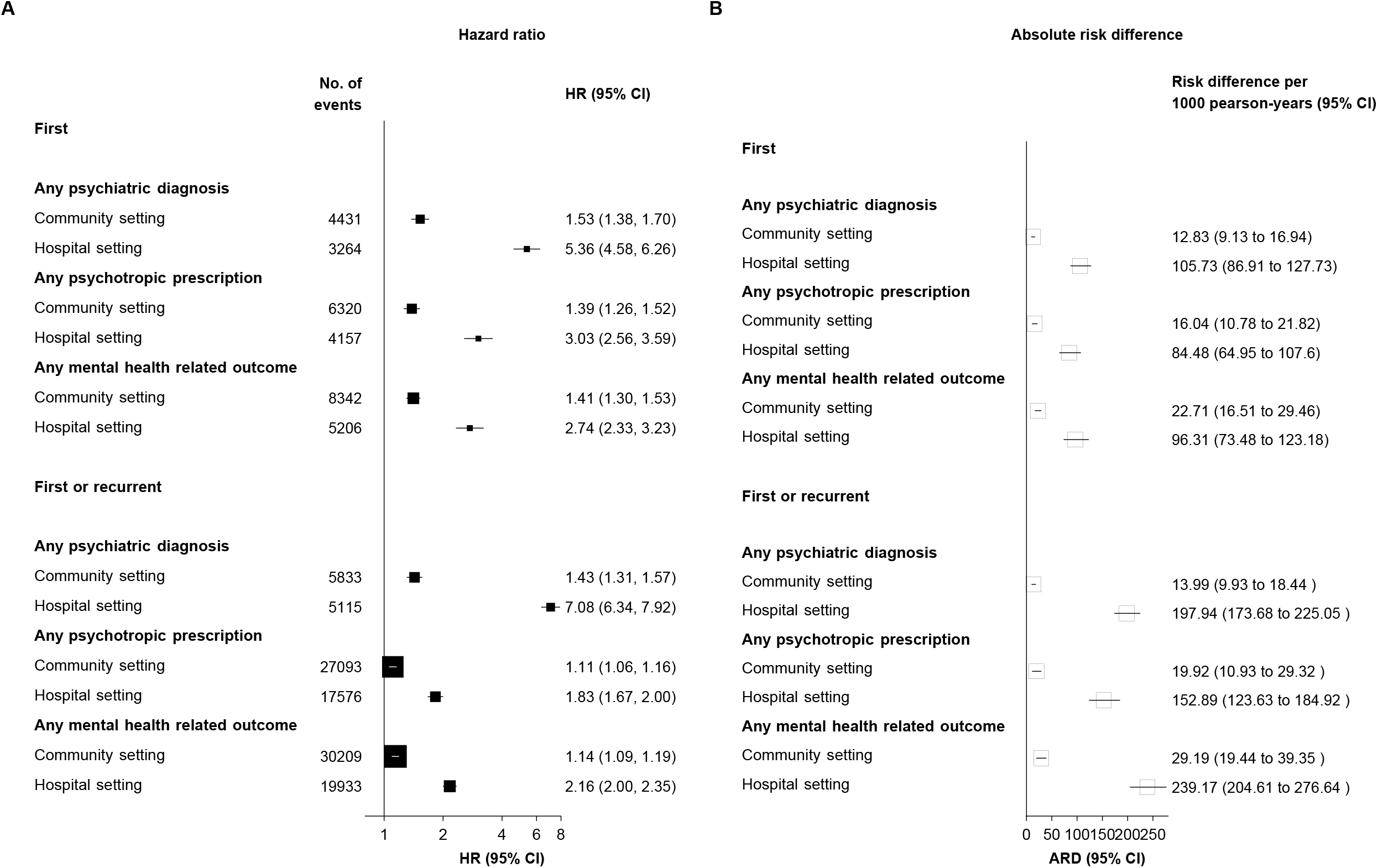
Risks of psychiatric diagnoses and prescriptions for psychotropic medications by test setting of participants in SARS-Cov-2 infection group compared with the contemporary control group. Mental health outcomes were ascertained after the SARS-CoV-2 infection until the end of follow-up. Hazard ratios were adjusted for predefined and data-driven covariates.

#### COVID-19 group versus historical control

After weighting, all characteristics were well balanced between infection and historical control groups (ASMD <0.1) and the distribution of index dates were fully aligned. The results were consistent with analyses using contemporary control as the referent group and showed increased risk of mental health outcomes in infection group compared with historical control group (**Supplementary Figure 3-5**).

#### *COVID-19 group versus other respiratory tract infection or* test-negative control

We assessed the risk of mental health outcomes in infection group compared with two additional control groups (respiratory tract infection, n=30598; test-negative, n=124806). Characteristics between groups were balanced after weighting. Compared with those with any respiratory tract infection, the risks of first mental health outcomes were decreased in infection group (**Supplementary Table 5**). Compared with test-negative control, the risks of first mental health outcomes were not significant or decreased in infection group (**Supplementary Table 5**).

### Sensitivity analyses

The results of main results of first mental health outcomes were robust in multiple sensitivity analyses. Sensitivity analyses using PS weighting, extending the look-back window for the data-driven covariates to three years, or excluding participants with history of outcomes in two years before follow-up indicated consistent results of increased risk of first mental health outcomes in infection group compared with contemporary control groups (**Supplementary Table 6**).

## Discussion

In this large-scale prospective population-based cohort, participants with SARS-CoV-2 infection were at increased risks of subsequent first psychiatric diagnoses including psychotic disorders, mood disorders, anxiety disorders, substance use disorders, and sleep disorders, and related prescriptions for psychotropic medications after COVID-19 compared with participants with no evidence of infection in the contemporary control group who experienced the same social and environmental stressors related to the COVID-19 pandemic. The results were consistent when comparing the SARS-CoV-2 infection group with the historical control group that predated the pandemic. These risks were evident even in those who tested positive in the community setting, likely consisting of infected people with mild or asymptomatic symptoms of COVID-19, and were highest in those who tested positive in the hospital and may have more severe COVID-19. Fully vaccinated participants with breakthrough infection were at lower risk of mental health outcomes compared with those with no or partial vaccination when they got infected. The increased risk of mental health outcomes was robust in multiple sensitivity analyses. Overall, these findings suggest that survivors of COVID-19 are at increased risk of subsequent psychiatric disorders and related psychotropic prescriptions. Vaccination may potentially have additional benefits of alleviating long-term psychiatric sequelae of COVID-19 beyond protecting against COVID-19 infection and severe complications.

Previous studies based on electronic health record (EHR) data suggested that individuals with or without a history of mental illness had an increased risk of psychiatric conditions in the following 3 to 12 months after acute infection of SARS-CoV-2 infection compared with individuals without evidence of the infection or individuals with other acute respiratory infections.^5-8^ Nonetheless, socioeconomic and lifestyle determinants associated with the SARS-CoV-2 infection and mental health outcomes and were largely unavailable in these studies, possibly leading to residual confounding. There may also be recording or surveillance bias due to restrictions and disruptions in patient help-seeking behaviors during the early period of the pandemic. In addition, the impact of vaccination status was not investigated in previous studies, and the association between vaccination and psychiatric sequelae of COVID-19 remain uncertain. Using a large prospective cohort recruited before the pandemic in the UK, our study provided more precise, representative risk estimates that corroborates previous EHR-based reports suggesting an increased risk of first psychiatric disorders as well as prescription for psychotropic medications after SARS-CoV-2 infection.

Notably, comparing test-positive with test-negative individuals or individuals with respiratory tract infections before the COVID-19 pandemic, we found no increased risk of mental health outcomes. Our results of test-negative control are consistent with a Danish registry study suggesting that the risks of mental health outcomes (depression, anxiety disorders, and psychosis) were not differential or even decreased in the infection group compared with the test-negative group.^19^ A UK primary care registry study also found that the risk of psychiatric morbidity in individuals with negative SARS-CoV-2 test results was increased compared with general population with no evidence of infection, and this association was similar to that observed in test-positive individuals.^20^ Although having a negative test result should not directly affect mental health outcomes, the testing behaviour in the circumstance of underlying non-infection is likely to indicate that unobserved confounders such as health anxiety, occupational and behavioral factors predict a higher risk of subsequent mental health outcomes. For example, healthcare workers who require more frequent COVID-19 tests even without any symptoms may experience excess psychological distress and be vulnerable to mental illness.^21,22^ In addition, individuals who seek a test could be experiencing health anxiety and are predisposed to mental health issues. A previous study has also suggested that individuals with negative test results had a higher proportion of prior mental health disorders than those with positive results.^20^ However, previous large US registry studies^6-8^ supporting an increased risk of mental health outcomes after COVID-19 did not specially compare risk between the test-positive and test negative groups, and this finding should be interpreted with caution and warrant further research. An alternative explanation to our findings could be that testing behavior rather than infection could at least partially account for the observed increase in mental health outcomes after COVID-19. However, our observations of a further increase in risk amongst those with severe (hospitalized) COVID-19, and in the unvaccinated or partially unvaccinated, would support a causal association between SARS-CoV-2 infection and mental health outcomes.

Our results of respiratory tract infections were consistent with two UK studies suggesting the increase in risk was larger for individuals with non-COVID-19 respiratory tract infections than individuals with SARS-CoV-2 infection.^5,20^ This is likely because individuals who were admitted to the hospital with respiratory tract infections may have a more severe condition, considering the majority of individuals in the SARS-CoV-2 infection group were tested positive in the community setting and were not admitted to the hospital. However, the US studies^6-8^ reported an increased risk of psychiatric sequelae after 6 to 12 months compared with the control group of influenza, which could be partly explained by the difference in study design (any respiratory tract infection in our study rather than only influenza), follow-up period and diagnostic practices in the US and UK. Of note, a recent US EHR study found that the increased risks of anxiety and mood disorders after SARS-CoV-2 infection were returned to baseline levels in the control group of other respiratory infections after 1-2 months, and the subsequent HRs from that time onwards were consistently lower than 1 over the 2-year follow-up.^23^

Our findings of the risk of psychiatric sequela by vaccination status also address the knowledge gap showing that vaccination (independent of vaccine type) potentially has additional benefits of alleviating long-term psychiatric sequelae of COVID-19 beyond protecting against COVID-19 infection and severe complications. A recent study in the UK suggested that long covid symptoms such as trouble sleeping, worry, and weakness were observed to decrease after vaccination and there was sustained improvement after two dose of vaccination.^24^ Given the existing large number of COVID-19 survivors (to date about 500 million globally) and the increasing infections worldwide accompanying the loosening of COVID-19 restrictions, the absolute risk of first incident psychiatric disorders may translate into a enormous global burden of mental health. Suppose the benefits of vaccination on long-term psychiatric manifestations of COVID-19 were confirmed by independent studies, the vaccine should be considered as part of public health strategies against the long-term symptoms of COVID-19, given the substantial medical costs associated with treating these related mental disorders. Policy makers and health systems should also develop priorities and long-term strategies for early identification and treatment of affected individuals to mitigate the psychiatric sequela and empower wellbeing especially in vulnerable survivors of COVID-19.

SARS-CoV-2 infection may impact subsequent mental health directly and indirectly through several plausible mechanisms at both biological and environmental levels. The COVID-19 pandemic and related public health interventions such as quarantine and social distancing may have long-term adverse mental health consequences, especially on vulnerable groups including patients diagnosed with COVID-19 and those with pre-existing mental disorders.^22,25^ These disproportional impacts may partly explain the increased risk of mental health outcomes after infection compared with the contemporary control, although participants in both groups experienced similar pandemic-related socioeconomic and environmental stressors. In addition, possible changes in behaviours such as decreased physical activity, having a poor diet, and increased avoidance of health care and social contact in some individuals following recovery from acute COVID-19 may also contribute to the increased risk of long-term psychiatric sequelae.^2,25^ Overlapping biological factors between viral infection and psychiatric disorders may also be implicated. Several possible underlying mechanisms include increased blood-brain barrier permeability and the central nervous system infiltration of SARS-CoV-2, chronic systemic immuno-inflammatory responses, dysregulation of microglia and astrocytes, and disturbances in synaptic signaling of upper layer excitatory neurons.^26,27^ Future studies are needed to explore whether post-COVID-19 psychiatric disorders result from SARS-CoV-2 infection itself, the disproportional adverse effects of pandemic-related factors, or a combination of both.

To our knowledge, this is the most extensive study to systematically explore the psychiatric sequelae of COVID-19 in a prospective cohort with comprehensive and reliable recorded data including socioeconomic and lifestyle factors and vaccination status that were largely unavailable in previous EHR-based studies. The potential benefits of vaccination on psychiatric sequelae reinforce the need for vaccination and support the ongoing global vaccination campaigns. Overall, the findings are robust given the large sample size, the use of PS weighting, and the consistent results in sensitivity and secondary analyses. However, the findings from this study should be interpreted with caution in the context of its limitations. First, concerns have been raised that participants in UK Biobank may be suboptimally representative of the whole population in the UK and were likely to be older and generally healthier. These issues primarily affect the estimates of absolute incidence rates. Despite the relative risk between comparison groups was largely not influenced, this might still limit the generalizability of our findings to other younger populations. Second, although we used robust statistical approaches such as PS weighting and PS matching based on a set of covariates to adjusted for the potential differences in characteristics between comparison groups, residual confounding cannot be ruled out in this observational study. Mendelian randomization (MR) is less susceptible to potential bias that are common in conventional observational studies. However, the heritability of 7 single nucleotide polymorphisms reached genome-wide significance identified in the current largest GWAS of SARS-CoV-2 infection was low at 0.17%,^28^ which may lead to weak instrument bias and preclude the conducting of valid MR analyses at the current stage. Future MR study are needed to further clarify whether the observed association are causal when robust genetic variants associated with COVID-19 are available. Third, a proportion of participants in the contemporary control group may have undiagnosed or untested COVID-19. However, this tends to make the risk estimates underestimate and thus lead to more conservative results. The linkage of UK Biobank participants to official national databases for COVID-19 testing and hospitalization meant that the likelihood of misclassification of infected and uninfected participants was minimized. Fourth, we did not statistically correct for multiple comparisons, although most results were significant at p-value less than 0.0001. Fifth, although we observed an significantly increased risk of a series of psychiatric diagnoses, the case number of several disorders was relatively small, limiting the further analysis of subcategories, especially severe ones such as schizophrenia. Finally, the risk estimates from our analyses may be representative of the mixed effect of several SARS-CoV-2 strains (the alpha and delta variants were dominant during different periods of follow-up), which should be cautiously extrapolated to novel variants, such as omicron. However, one recent study found the risks of neurological and psychiatric outcomes after the emergence of the omicron (B.1.1.529) variant were similar to these after the delta (B.1.617.2) variant.^23^ The epidemiology of COVID-19 psychiatric sequela may also change with the evolving pandemic, emerging variants, and increasing vaccine uptake and further studies are warranted.

In conclusion, in this large-scale prospective cohort study, people who survived the acute phase of COVID-19 were at increased risk of subsequent first psychiatric disorders and psychotropic prescriptions. These risks was significantly reduced in fully vaccinated people with breakthrough infection compared to those with unvaccinated or partially vaccinated people with non-breakthrough infection. Future independent studies are needed to verify the potential benefits of the vaccine on the psychiatric sequela of COVID-19 and to inform other approaches to empower mental health wellbeing. Identification and treatment of psychiatric disorders among survivors of SARS-CoV-2 infection should be a priority in the long-term management of COVID-19.

## Data Availability

Bonafide researchers can apply to use the UK Biobank dataset by registering and applying at https://ukbiobank.ac.uk/register-apply/. Any additional summary data generated and/or analyzed during the current study are available from the corresponding author on reasonable request.

## Declaration of interests

### Author Contributions

Drs Wang and Xie had full access to all the data in the study and take responsibility for the integrity of the data and the accuracy of the data analyses. *Concept and design*: Wang, Xie, and Prieto-Alhambra. *Acquisition, analysis, or interpretation of data*: Wang, Xie, and Prieto-Alhambra. *Drafting of the manuscript*: Wang. *Critical revision of the manuscript for important intellectual content*: All authors. *Statistical analysis*: Xie. *Obtained funding*: Prieto-Alhambra. *Administrative, technical, or material support*: Prieto-Alhambra. *Supervision*: Prieto-Alhambra.

### Ethical approval

All participants provided written informed consent at the UKBB cohort recruitment. This study received ethical approval from UKBB Ethics Advisory Committee (EAC) and was performed under the application of 65397.

