## Supplementary tables and figures for "Long-term mental health outcomes after SARS-CoV-2 infection: prospective cohort study"


| Table of Contents | Page |
|---|---|
| **SUPPLEMENTARY FIGURES** | |
| Supplementary Figure 1. Distribution of SARS-CoV-2 infections in the UK Biobank over the study period | **1** |
| Supplementary Figure 2. Risks of first or recurrent psychiatric diagnoses and prescriptions for psychotropic medications after SARS-Cov-2 infection compared with the contemporary control group | **2** |
| Supplementary Figure 3. Risks of first psychiatric diagnoses and prescriptions for psychotropic medications after SARS-CoV-2 infection compared with the historical control group | **3** |
| Supplementary Figure 4. Risks of first or recurrent psychiatric diagnoses and prescriptions for psychotropic medications after SARS-CoV-2 infection compared with the historical control group | **4** |
| Supplementary Figure 5. Risks of composite mental health outcomes after SARS-CoV-2 infection compared with the historical control group | **5** |
| **SUPPLEMENTARY TABLES** | |
| Supplementary Table 1. Definition of mental health related outcomes | **6** |
| Supplementary Table 2. Demographic and medical characteristics of SARS-CoV-2 infection, contemporary control, and historical control cohorts before weighting | **8** |
| Supplementary Table 3. Risks of first composite mental health outcomes in participants with breakthrough infection compared with non-breakthrough infection | **10** |
| Supplementary Table 4. Risks of first composite mental health outcomes in participants who tested positive in hospital setting compared with those who tested positive in community setting | **11** |
| Supplementary Table 5. Risks of first composite mental health outcomes in SARS-CoV-2 infection group compared with the control groups of respiratory tract infection or the test-negative control groups | **12** |
| Supplementary Table 6. Sensitivity analyses for the risks of first composite mental health outcomes in infection group compared with contemporary control groups | **13** |

#### Supplementary Figure 1. Distribution of SARS-CoV-2 infections in the UK Biobank over the study period

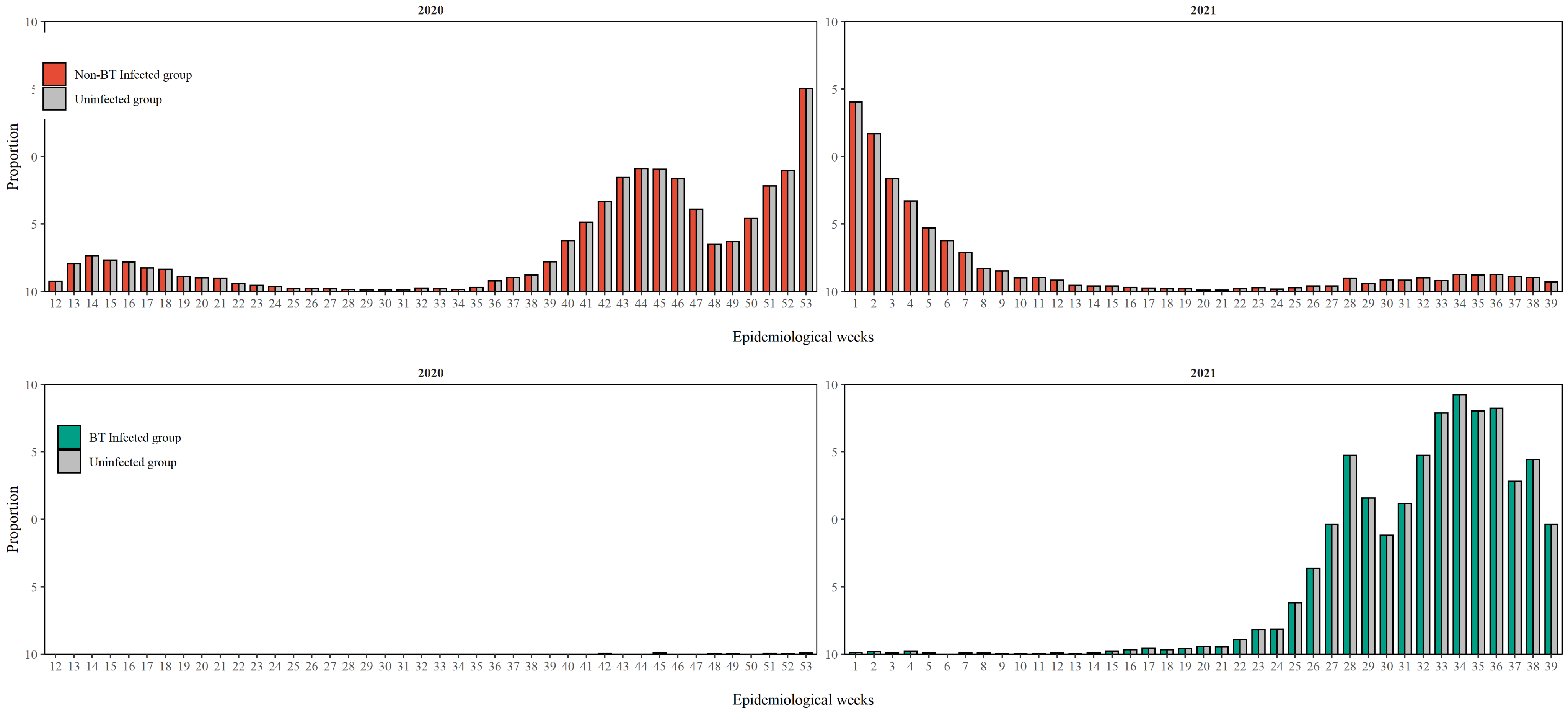

**Figure S2. Risks of first or recurrent psychiatric diagnoses and prescriptions for psychotropic medications after SARS-CoV-2 infection compared with the contemporary control group**

**A** — Hazard ratio

**B** — Absolute risk difference

| Outcome | No. of events | HR (95% CI) | Risk difference per 1000 pearson-years (95% CI) |
|---|---|---|---|
| **Psychiatric diagnoses** | | | |
| Psychotic disorders | 199 | 3.22 (2.16, 4.80) | 1.67 (0.87 to 2.87) |
| Mood disorders | 3212 | 2.43 (2.18, 2.71) | 18.59 (15.38 to 22.17) |
| Depressive episode | 3023 | 2.40 (2.15, 2.68) | 17.13 (14.06 to 20.56) |
| Mania/Bipolar affective disorder | 235 | 2.84 (1.91, 4.24) | 1.74 (0.85 to 3.06) |
| Anxiety disorders | 2889 | 2.20 (1.96, 2.47) | 14.15 (11.30 to 17.36) |
| Phobic anxiety disorder | 181 | 1.74 (1.02, 2.97) | 0.57 (0.01 to 1.53) |
| Panic disorder | 163 | 2.12 (1.26, 3.58) | 0.76 (0.18 to 1.76) |
| Generalized anxiety disorder | 2644 | 2.24 (1.98, 2.53) | 13.35 (10.60 to 16.46) |
| PTSD | 78 | 3.26 (1.83, 5.80) | 0.67 (0.25 to 1.42) |
| Substance use disorders | 2330 | 1.79 (1.55, 2.07) | 7.75 (5.40 to 10.46) |
| Alcohol use disorder | 663 | 2.51 (1.97, 3.19) | 4.02 (2.59 to 5.84) |
| Tobacco use disorder | 1792 | 1.52 (1.28, 1.80) | 3.96 (2.12 to 6.14) |
| Sleep disorders | 1348 | 1.98 (1.68, 2.33) | 5.38 (3.75 to 7.29) |
| **Psychotropic medications** | | | |
| Antipsychotics | 2233 | 1.78 (1.54, 2.07) | 7.42 (5.12 to 10.10) |
| Antidepressants | 17350 | 1.11 (1.04, 1.17) | 8.39 (3.36 to 13.73) |
| SSRI | 15578 | 1.11 (1.04, 1.18) | 7.43 (2.69 to 12.48) |
| SNRI | 1877 | 1.13 (0.94, 1.34) | 1.03 (-0.47 to 2.81) |
| Other antidepressants | 6181 | 1.38 (1.25, 1.52) | 10.26 (6.80 to 14.08) |
| Benzodiazepines | 5716 | 1.44 (1.30, 1.59) | 10.79 (7.34 to 14.62) |
| Opioids | 13843 | 1.19 (1.11, 1.28) | 11.90 (6.94 to 17.21) |

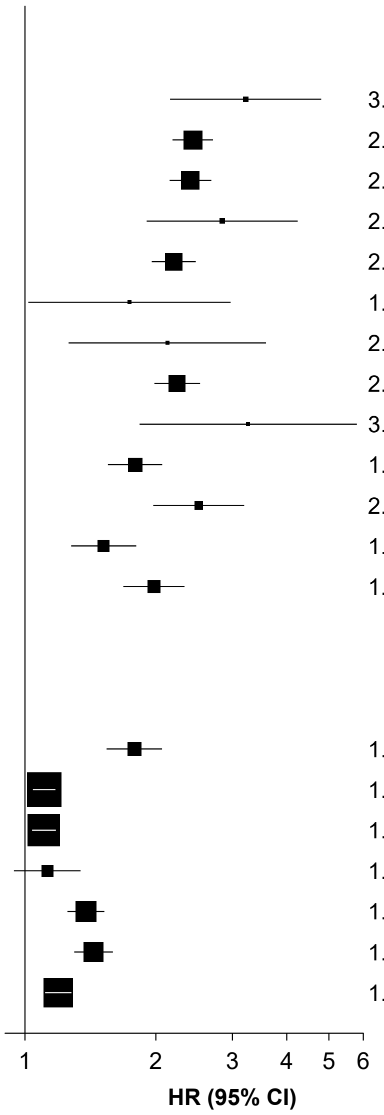


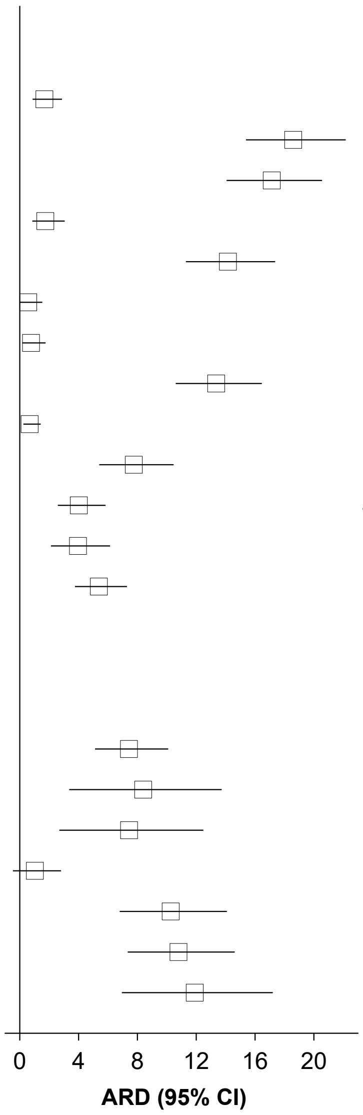


Mental health outcomes were ascertained after the SARS-CoV-2 infection until the end of follow-up. Hazard ratios were adjusted for predefined and data-driven covariates. SSRI=selective serotonin reuptake inhibitor; SNRI =serotonin-noradrenaline reuptake inhibitor.

**Figure S3. Risks of first psychiatric diagnoses and prescriptions for psychotropic medications after SARS-CoV-2 infection compared with the historical control group**

| | A: Hazard ratio — No. of events | A: HR (95% CI) | B: Absolute risk difference — Risk difference per 1000 pearson-years (95% CI) |
|---|---|---|---|
| **Psychiatric diagnoses** | | | |
| Psychotic disorders | 116 | 3.08 (1.61, 5.87) | 0.91 (0.27 to 2.13) |
| Mood disorders | 2061 | 2.91 (2.52, 3.36) | 15.59 (12.39 to 19.29) |
| Depressive episode | 1954 | 2.94 (2.54, 3.40) | 14.91 (11.84 to 18.46) |
| Mania/Bipolar affective disorder | 145 | 3.10 (1.73, 5.57) | 1.25 (0.43 to 2.72) |
| Anxiety disorders | 1937 | 2.96 (2.54, 3.43) | 15.00 (11.85 to 18.67) |
| Phobic anxiety disorder | 126 | 3.03 (1.63, 5.61) | 1.02 (0.32 to 2.32) |
| Panic disorder | 126 | 3.51 (1.96, 6.28) | 1.27 (0.49 to 2.68) |
| Generalized anxiety disorder | 1771 | 3.04 (2.61, 3.55) | 14.26 (11.21 to 17.81) |
| PTSD | 47 | 4.57 (2.08, 10.04) | 0.60 (0.18 to 1.52) |
| Substance use disorders | 1687 | 1.85 (1.53, 2.25) | 6.03 (3.73 to 8.83) |
| Alcohol use disorder | 530 | 2.73 (2.02, 3.69) | 3.67 (2.16 to 5.70) |
| Tobacco use disorder | 1279 | 1.57 (1.24, 1.98) | 3.08 (1.28 to 5.35) |
| Sleep disorders | 720 | 3.14 (2.50, 3.93) | 5.89 (4.14 to 8.08) |
| **Psychotropic medications** | | | |
| Antipsychotics | 324 | 2.81 (1.97, 3.99) | 2.25 (1.21 to 3.73) |
| Antidepressants | 1779 | 1.61 (1.33, 1.95) | 4.86 (2.62 to 7.57) |
| SSRI | 1451 | 1.68 (1.36, 2.07) | 4.36 (2.29 to 6.91) |
| SNRI | 444 | 1.46 (1.03, 2.09) | 0.87 (0.05 to 2.03) |
| Other antidepressants | 1467 | 2.22 (1.84, 2.68) | 7.39 (5.08 to 10.17) |
| Benzodiazepines | 2528 | 1.83 (1.56, 2.15) | 8.96 (6.05 to 12.37) |
| Opioids | 6048 | 1.39 (1.24, 1.55) | 10.62 (6.64 to 15.06) |

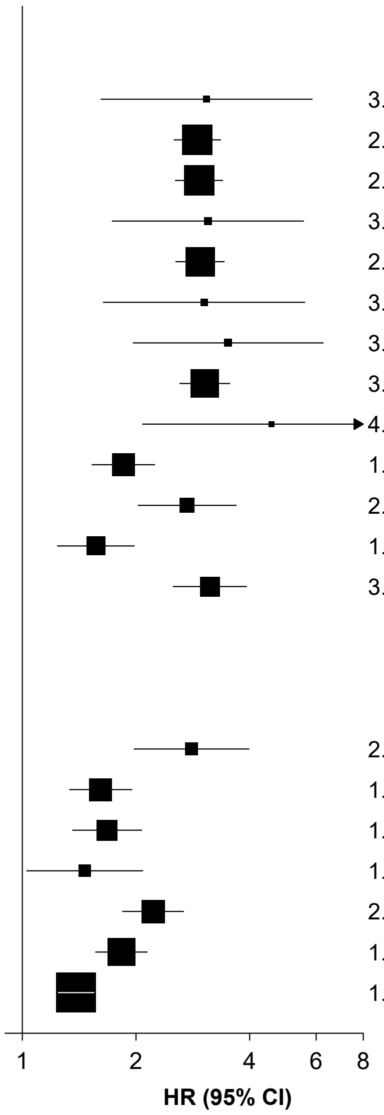


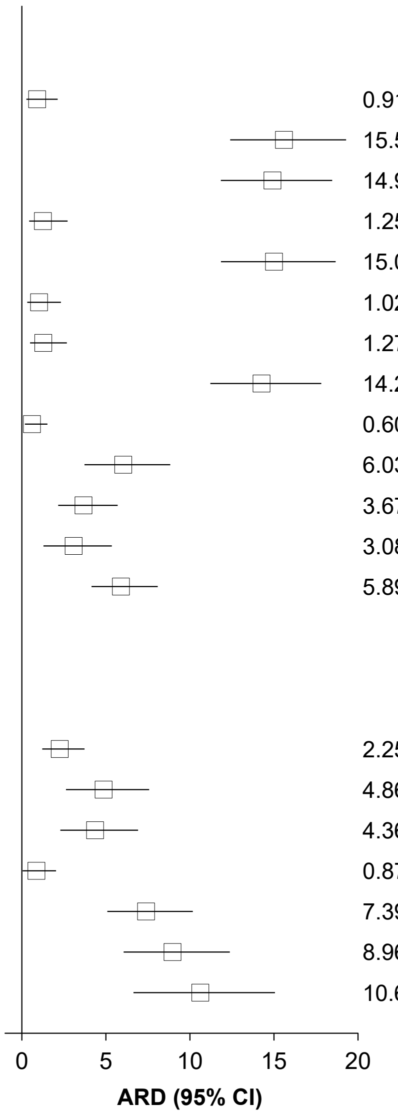


Mental health outcomes were ascertained after the SARS-CoV-2 infection until the end of follow-up. Hazard ratios were adjusted for predefined and data-driven covariates. SSRI=selective serotonin reuptake inhibitor; SNRI =serotonin-noradrenaline reuptake inhibitor.

**Figure S4. Risks of first or recurrent psychiatric diagnoses and prescriptions for psychotropic medications after SARS-CoV-2 infection compared with the historical control group**

**A** — **Hazard ratio**

**B** — **Absolute risk difference**

| | No. of events | HR (95% CI) | Risk difference per 1000 pearson-years (95% CI) |
|---|---|---|---|
| **Psychiatric diagnoses** | | | |
| Psychotic disorders | 200 | 3.42 (2.20, 5.31) | 1.80 (0.89 to 3.21) |
| Mood disorders | 2915 | 3.10 (2.75, 3.49) | 23.80 (19.84 to 28.26) |
| Depressive episode | 2713 | 3.09 (2.73, 3.49) | 21.99 (18.23 to 26.24) |
| Mania/Bipolar affective disorder | 230 | 3.56 (2.30, 5.51) | 2.30 (1.17 to 4.06) |
| Anxiety disorders | 2494 | 3.06 (2.69, 3.49) | 20.10 (16.47 to 24.25) |
| Phobic anxiety disorder | 148 | 2.74 (1.52, 4.93) | 1.04 (0.31 to 2.35) |
| Panic disorder | 152 | 3.02 (1.73, 5.26) | 1.25 (0.45 to 2.63) |
| Generalized anxiety disorder | 2278 | 3.15 (2.75, 3.60) | 19.05 (15.53 to 23.07) |
| PTSD | 74 | 4.42 (2.34, 8.34) | 0.93 (0.36 to 1.99) |
| Substance use disorders | 2394 | 1.95 (1.67, 2.29) | 9.46 (6.62 to 12.79) |
| Alcohol use disorder | 702 | 2.73 (2.10, 3.54) | 4.84 (3.08 to 7.12) |
| Tobacco use disorder | 1834 | 1.64 (1.36, 1.99) | 4.95 (2.74 to 7.64) |
| Sleep disorders | 1186 | 2.79 (2.33, 3.34) | 8.23 (6.10 to 10.78) |
| **Psychotropic medications** | | | |
| Antipsychotics | 2244 | 1.92 (1.62, 2.26) | 8.53 (5.81 to 11.75) |
| Antidepressants | 18563 | 1.16 (1.08, 1.24) | 13.06 (6.79 to 19.76) |
| SSRI | 16814 | 1.16 (1.08, 1.24) | 11.72 (5.75 to 18.13) |
| SNRI | 1870 | 1.17 (0.96, 1.42) | 1.36 (-0.30 to 3.37) |
| Other antidepressants | 6078 | 1.46 (1.31, 1.63) | 12.00 (8.07 to 16.39) |
| Benzodiazepines | 6064 | 1.41 (1.25, 1.58) | 10.49 (6.57 to 14.89) |
| Opioids | 14627 | 1.18 (1.09, 1.28) | 11.65 (6.02 to 17.74) |

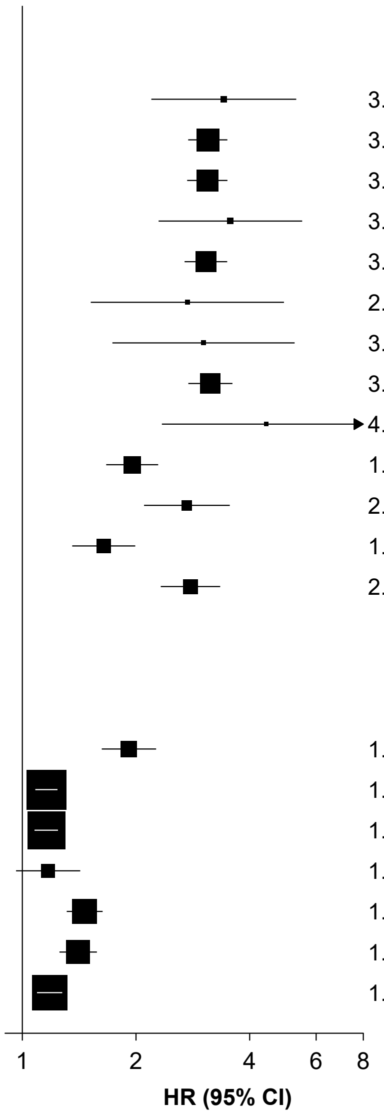


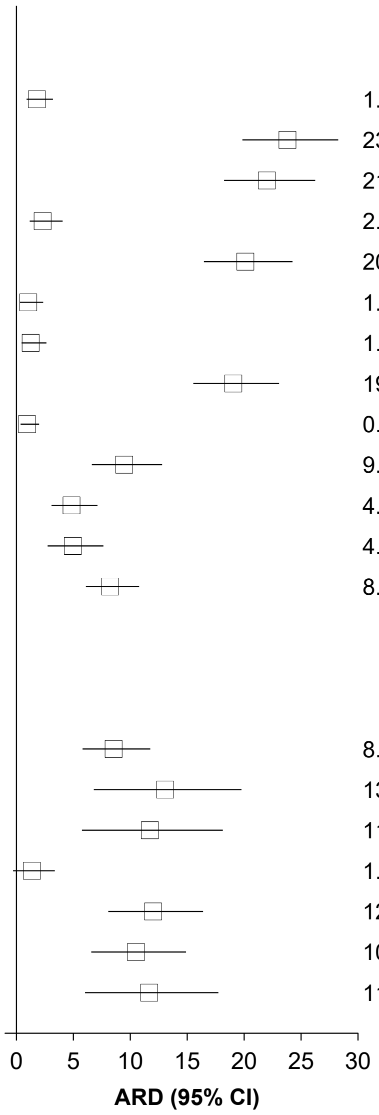


Mental health outcomes were ascertained after the SARS-CoV-2 infection until the end of follow-up. Hazard ratios were adjusted for predefined and data-driven covariates. SSRI=selective serotonin reuptake inhibitor; SNRI =serotonin-noradrenaline reuptake inhibitor.

**Figure S5. Risks of composite mental health outcomes after SARS-CoV-2 infection compared with the historical control group**

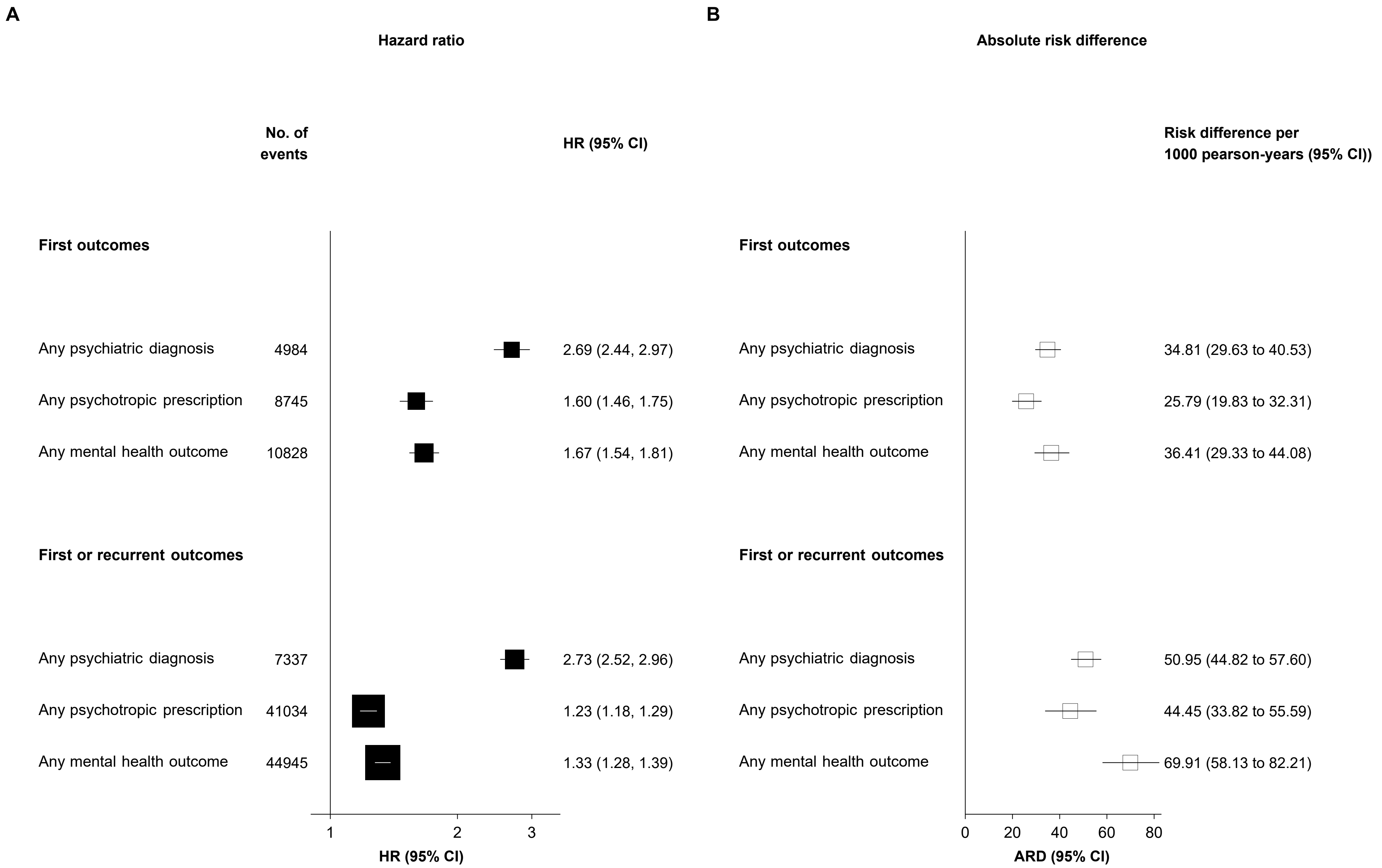


Mental health outcomes were ascertained after the SARS-CoV-2 infection until the end of follow-up. Hazard ratios were adjusted for predefined and data-driven covariates.

**Supplementary Table 1. Definition of mental health related outcomes**

| Outcome | Type of definition | Definition |
|---|---|---|
| **Psychiatric diagnoses** | | |
| Psychotic disorders | ICD-10 | F20-F29 |
| Mood disorders | ICD-10 | F30-F39 |
| Mania/Bipolar affective disorder | ICD-10 | F30-F31 |
| Depressive episode | ICD-10 | F32 |
| Anxiety disorders | ICD-10 | F40-F48 |
| Phobic anxiety disorders | ICD-10 | F40 |
| Panic disorder | ICD-10 | F41.0 |
| Generalized anxiety disorder | ICD-10 | F41.1 |
| Posttraumatic stress disorder | ICD-10 | F43.1 |
| Substance use disorders | ICD-10 | F10-F19 |
| Illicit drug use disorder | ICD-10 | F12, F14-F16, F18-F19 |
| Alcohol use disorder | ICD-10 | F10 |
| Tobacco use disorder | ICD-10 | F17 |
| Sedative/hypnotics use disorder | ICD-10 | F13 |
| Sleep disorders | ICD-10 | F51, G47 |
| **Prescriptions for psychotropic medications** | | |
| Antipsychotics | Medication | Amisulpride, Aripiprazole, Clozapine, Olanzapine, Paliperidone, Quetiapine, Risperidone |
| Antidepressant | Medication | Any SSRI, SNRI, or other antidepressant medications |
| SSRI | Medication | Citalopram, Escitalopram, Dapoxetine, Escitalopram, Fluoxetine, Fluvoxamine, Paroxetine, Sertraline, Vortioxetine, Trazodone |
| SNRI | Medication | Venlafaxine, Duloxetine, Milnacipran |
| Other antidepressant drugs | Medication | Flupentixol, Doxepin, Bupropion, Amoxapine, Maprotiline, Mianserin, Mirtazapine |
| Benzodiazepines | Medication | Diazepam, Nitrazepam, Lorazepam, Flurazepam, Temazepam, Flunitrazepam, Alprazolam, Clobazam, Clonazepam, Chlordiazepoxide, Lormetazepam, Oxazepam, Midazolam |
| Opioids | | |
| Opioid prescription | Medication | Codeine, Hydromorphone, Morphine, Opium, Oxycodone, Alfentanil, Dihydrocodeine, |

| | | |
|---|---|---|
| | | Diphenoxylate, Fentanyl, Remifentanil, Pentazocine, Tramadol, Nalbuphine |
| Opioid use disorder | ICD-10 | F11 |
| Naloxone or naltrexone | Medication | Naloxone or naltrexone |
| Methadone | Medication | Methadone |
| Buprenorphine | Medication | Buprenorphine |
| **Composite outcomes** | | |
| Any psychiatric diagnosis | Composite | Any ICD-10 code listed |
| Any prescriptions for psychotropic medications | Composite | Any psychotic medication listed |
| Any mental health related conditions | Composite | Any ICD-10 code or psychotic medication listed |

**Supplementary Table 2. Demographic and medical characteristics of SARS-CoV-2 infection, contemporary control, and historical control cohorts before weighting**

| Characteristics | SARS-Cov-2 infection (n=26181) | Contemporary control (n=380398) | Historical control (n=384030) | ASMD between infection and contemporary control* | ASMD between infection and historical control* |
|---|---|---|---|---|---|
| Age, mean (sd) | 66.0 (8.5) | 68.8 (8.1) | 67.87 (8.06) | 0.34 | 0.23 |
| Sex, male (%) | 12340 (47.1) | 169377 (44.5) | 171447 (44.6) | 0.05 | 0.05 |
| Ethnicity, White (%) | 22151 (84.6) | 356546 (93.7) | 360042 (93.8) | 0.30 | 0.30 |
| Index of Multiple Deprivation, mean (sd) | 20.5 (14.9) | 17.3 (13.7) | 17.3 (13.75) | 0.22 | 0.22 |
| Body Mass Index, mean (sd) | 28.1 (5.0) | 27.3 (4.7) | 27.3 (4.71) | 0.17 | 0.16 |
| Current smoker (%) | 2843 (10.9) | 36704 (9.6) | 37312 (9.7) | 0.01 | 0.23 |
| Current drinker (%) | 23771 (90.8) | 349770 (91.9) | 352987 (91.9) | 0.11 | 0.05 |
| Physical activity, high level (%)[#] | 8554 (32.7) | 124637 (32.8) | 125638 (32.7) | 0.05 | 0.06 |
| Vaccination status, fully-vaccinated (%) | 9739 (37.2) | 149496 (39.3) | NA | 0.04 | NA |
| Medications (%)[†] | | | | | |
| Lipid lowering drugs | 8704 (33.2) | 135512 (35.6) | 132446 (34.5) | 0.05 | 0.03 |
| RAS inhibitors | 6096 (23.3) | 91022 (23.9) | 90264 (23.5) | 0.02 | 0.01 |
| Other anti-hypertensives | 2882 (11.0) | 41046 (10.8) | 41577 (10.8) | 0.01 | 0.01 |
| Anticoagulants | 1120 (4.3) | 16278 (4.3) | 15207 (4.0) | 0.01 | 0.02 |
| Antiplatelet drugs | 3030 (11.6) | 42694 (11.2) | 42483 (11.1) | 0.01 | 0.02 |
| Proton pump inhibitors | 8491 (32.4) | 110972 (29.2) | 107396 (28.0) | 0.07 | 0.10 |
| Diabetes medicines | 2235 (8.5) | 25491 (6.7) | 25392 (6.6) | 0.07 | 0.07 |
| Systemic glucocorticoids | 1746 (6.7) | 19850 (5.2) | 24417 (6.4) | 0.06 | 0.01 |
| Immunosuppressants | 341 (1.3) | 4734 (1.2) | 4772 (1.2) | 0.01 | 0.01 |
| Antineoplastic agents | 36 (0.1) | 437 (0.1) | 407 (0.1) | 0.01 | 0.01 |
| Coexisting conditions (%)[†] | | | | | |
| Acquired immunodeficiency syndrome | 27 (0.1) | 394 (0.1) | 384 (0.1) | 0.01 | 0.01 |
| Cancer | 2419 (9.2) | 40851 (10.7) | 39391 (10.3) | 0.05 | 0.03 |
| Cerebrovascular disease | 625 (2.4) | 8811 (2.3) | 8289 (2.2) | 0.01 | 0.02 |
| Chronic obstructive pulmonary disease | 4807 (18.4) | 62961 (16.6) | 62416 (16.3) | 0.05 | 0.06 |
| Chronic kidney disease | 1399 (5.3) | 20033 (5.3) | 18945 (4.9) | 0.01 | 0.02 |

| | | | | | |
|---|---|---|---|---|---|
| Congestive heart failure | 506 (1.9) | 5659 (1.5) | 5066 (1.3) | 0.03 | 0.05 |
| Dementia | 485 (1.9) | 3025 (0.8) | 2551 (0.7) | 0.09 | 0.11 |
| Diabetes (uncomplicated) | 2952 (11.3) | 36070 (9.5) | 34821 (9.1) | 0.06 | 0.07 |
| Diabetes (end-organ damage) | 961 (3.7) | 11413 (3.0) | 10927 (2.8) | 0.04 | 0.05 |
| Hemiplegia | 36 (0.1) | 415 (0.1) | 417 (0.1) | 0.01 | 0.01 |
| Liver disease | 220 (0.8) | 2823 (0.7) | 2750 (0.7) | 0.01 | 0.01 |
| Peptic ulcer | 703 (2.7) | 8900 (2.3) | 8855 (2.3) | 0.02 | 0.02 |
| Rheumatoid arthritis | 818 (3.1) | 11262 (3.0) | 10712 (2.8) | 0.01 | 0.02 |
| Blood pressure, mean (sd), mm Hg | | | | | |
| Systolic blood pressure | 82.1 (10.6) | 82.1 (10.5) | 82.1 (10.5) | 0.01 | 0.01 |
| Diastolic blood pressure | 137.4 (18.8) | 139.5 (19.3) | 139.5 (19.3) | 0.11 | 0.11 |
| Hospital admissions, mean (sd) † | 0.58 (2.79) | 0.37 (1.85) | 0.43 (2.02) | 0.09 | 0.06 |

Abbreviations: SD, standard deviation; MET, metabolic equivalent of task; ASMD, absolute standardized mean difference.

### Physical activity status was measured by the International Physical Activity Questionnaire (IPAQ).

†Data collected within past one year of $T_0$ from primary care records.

*ASMD ≤0.10 is considered good balance between comparison groups.

**Supplementary Table 3. Risks of first composite mental health outcomes in participants with breakthrough infection compared with non-breakthrough infection**

| Outcome | Hazard ratio (95% CI) | Incidence rate per 1000 people at one year (95% CI) | | Difference in incidence rate per 1000 people at one year (95% CI) |
|---|---|---|---|---|
| | | Breakthrough infection | Non-breakthrough infection | |
| Any psychiatric diagnosis | 0.26 (0.18-0.36) | 40.95 (38.01-44.05) | 51.60 (50.55-52.68) | - |
| Any psychotropic prescription | 0.47 (0.33-0.66) | 45.51 (42.23-48.98) | 69.13 (67.83-70.44) | - |
| Any mental health related outcome | 0.41 (0.30-0.56) | 64.49 (60.55-68.63) | 90.35 (88.84-91.87) | - |

Mental health outcomes were ascertained after the SARS-CoV-2 infection until the end of follow-up. Hazard ratios were adjusted for predefined and data-driven covariates.

**Supplementary Table 4. Risks of first composite mental health outcomes in participants who tested positive in hospital setting compared with those who tested positive in community setting**

| Outcome | Hazard ratio (95% CI) | Incidence rate per 1000 people at one year (95% CI) | | Difference in incidence rate per 1000 people at one year (95% CI) |
|---|---|---|---|---|
| | | Hospital setting | Community setting | |
| Any psychiatric diagnosis | 3.60 (2.98-4.45) | 133.14 (128.65-137.74) | 38.41 (37.48-39.36) | - |
| Any psychotropic prescription | 2.22 (1.82-2.67) | 124.97 (120.44-129.63) | 58.00 (56.78-59.24) | - |
| Any mental health related outcome | 1.96 (1.64-2.34) | 151.00 (145.80-156.34) | 78.90 (77.47-80.36) | - |

Mental health outcomes were ascertained after the SARS-CoV-2 infection until the end of follow-up. Hazard ratios were adjusted for predefined and data-driven covariates.

**Supplementary Table 5. Risks of first composite mental health outcomes in SARS-CoV-2 infection group compared with the control groups of respiratory tract infection or the test-negative control groups**

| Control groups | Any first psychiatric diagnoses (95% CI) | | Any psychotropic prescriptions (95%) | | Any mental health related outcomes (95% CI) | |
|---|---|---|---|---|---|---|
| | HR | ARD | HR | ARD | HR | ARD |
| Any respiratory tract infection | 0.79 (0.67-0.93) | -13.13 (-20.60 to -4.33) | 0.79 (0.68-0.92) | -17.70 (-27.39 to -6.35) | 0.74 (0.64-0.84) | -31.61 (-42.91 to -18.64) |
| Test-negative | 0.67 (0.61-0.73) | -24.48 (-28.69 to -19.87) | 0.94 (0.86-1.02) | -4.64 (-10.24 to 1.47) | 0.75 (0.70-0.81) | -29.01 (-35.47 to -22.03) |

**Supplementary Table 6. Sensitivity analyses for the risks of first composite mental health outcomes in infection group compared with contemporary control groups**

| Analysis | Any first psychiatric diagnoses (95% CI) | | Any psychotropic prescriptions (95%) | | Any mental health related outcomes (95% CI) | |
|---|---|---|---|---|---|---|
| | HR | ARD | HR | ARD | HR | ARD |
| Using PS matching | 1.87 (1.72-2.04) | - | 1.49 (1.37-1.62) | - | 1.48 (1.37-1.59) | - |
| Using data-driven covariates within three years | 2.02 (1.85-2.21) | 24.87 (20.71 to 29.40) | 1.61 (1.48-1.75) | 25.41 (20.00 to 31.29) | 1.58 (1.47-1.70) | 32.18 (25.89 to 38.96) |
| Excluding participants with history in two years before follow-up | 2.04 (1.86-2.26) | 21.54 (17.63 to 25.86) | 1.69 (1.55 to 1.85) | 25.13 (19.81 to 30.96) | 1.66 (1.53 to 1.80) | 31.02 (24.84 to 37.72) |
